# *SETBP1* variants outside the degron disrupt DNA-binding and transcription independent of protein abundance to cause a heterogeneous neurodevelopmental disorder

**DOI:** 10.1101/2022.03.04.22271462

**Authors:** Maggie MK Wong, Rosalie A Kampen, Ruth O Braden, Gökberk Alagöz, Michael S Hildebrand, Christopher Barnett, Meghan Barnett, Alfredo Brusco, Diana Carli, Bert BA de Vries, Alexander JM Dingemans, Frances Elmslie, Giovanni B Ferrero, Nadieh A Jansen, Ingrid MBH van de Laar, Alice Moroni, David Mowat, Lucinda Murray, Francesca Novara, Angela Peron, Ingrid E Scheffer, Fabio Sirchia, Samantha J Turner, Aglaia Vignoli, Arianna Vino, Sacha Weber, Wendy K Chung, Marion Gerard, Vanessa López-González, Elizabeth Palmer, Angela T Morgan, Bregje W van Bon, Simon E Fisher

## Abstract

Germline *de novo SETBP1* variants cause clinically distinct and heterogeneous neurodevelopmental disorders. Heterozygous missense variants at a hotspot encoding a canonical degron lead to SETBP1 accumulation and Schinzel-Giedion syndrome (SGS), a rare severe developmental disorder involving multisystem malformations. Heterozygous loss-of-function variants result in *SETBP1* haploinsufficiency disorder which is phenotypically much milder than SGS. Following an initial description of four individuals with atypical SGS carrying heterozygous missense variants adjacent to the degron, a few individual cases of variants outside the degron were reported. Due to the lack of systematic investigation of genotype-phenotype associations of different types of *SETBP1* variants, and limited understanding of the roles of the gene in brain development, the extent of clinical heterogeneity and how this relates to underlying pathophysiological mechanisms remain elusive, imposing challenges for diagnosis and patient care. Here, we present a comprehensive investigation of the largest cohort to-date of individuals carrying *SETBP1* missense variants outside the degron (n=18, including one in-frame deletion). We performed thorough clinical and speech phenotyping with functional follow-up using cellular assays and transcriptomics. Our findings suggest that such variants cause a clinically and functionally variable developmental syndrome, showing only partial overlaps with classical SGS and *SETBP1* haploinsufficiency disorder, and primarily characterised by intellectual disability, epilepsy, speech and motor impairment. We provide evidence of loss-of-function pathophysiological mechanisms impairing ubiquitination, DNA-binding and transcription. In contrast to SGS and *SETBP1* haploinsufficiency, these effects are independent of protein abundance. Overall, our study provides important novel insights into diagnosis, patient care and aetiology of SETBP1-related disorders.

## Introduction

Different types of germline variants in the gene encoding SET binding protein 1 (*SETBP1*) (NM_015559.2) lead to distinct and phenotypically heterogeneous developmental syndromes^1^. Schinzel–Giedion syndrome (SGS, MIM #269150)^2^ is a rare and severe disorder with multisystem malformations that include recognizable facial characteristics, neurological problems (including severe intellectual disability, intractable epilepsy, cerebral blindness and deafness) and various congenital anomalies (such as heart defects, and genital, kidney and bone abnormalities)^3–6^. The majority of patients with a molecularly confirmed diagnosis have a shortened lifespan and often do not survive beyond the first decade^6^. In 2010, heterozygous germline *de novo* missense variants in one section of the *SETBP1* gene were identified as a cause of SGS. The variants cluster in a hotspot of 12 base pairs coding for four amino acids (residues 868 to 871) in the SKI domain of the SETBP1 protein^2,6^. These four amino acid residues are part of a canonical degron sequence recognised by ubiquitin E3 ligases and important for regulating protein degradation, and the variants are thought to cause SGS via gain-of-function mechanisms. Intriguingly, overlapping somatic *SETBP1* variants have been identified recurrently in several forms of myeloid leukaemia^6,7^. These overlapping somatic variants are more disruptive to the degron, implying a higher functional threshold to cause cancer^6^, and it is thought that germline variants might lead to features similar to cancer cells such as increased proliferation and accumulation in DNA damage but via different cell type-specific pathways^8^.

In contrast, *SETBP1*-specific deletions and *de novo* loss-of-function (LoF) point mutations disrupting the gene result in *SETBP1* haploinsufficiency disorder^9^, (MIM #616078), a syndrome that is phenotypically milder than SGS^10–14^. The heterozygous *SETBP1*-specific deletions lead to a loss of the locus while the LoF variants (nonsense and frameshift variants) are predicted to lead to nonsense-mediated decay of the mutated *SETBP1* transcript, both of which therefore result in reduced dosage of SETBP1 protein (haploinsufficiency). Unlike classical SGS, individuals carrying such variants do not show major congenital or growth anomalies. Our recent systematic gene-driven studies of a large cohort with confirmed *SETBP1* LoF variants revealed a far broader clinical severity spectrum than previously reported^15,16^. Despite subtle overlapping facial dysmorphisms, and in contrast to SGS, the affected individuals do not present with a recognizable facial gestalt or specific features of dysmorphisms. The main clinical features include moderate-to-severe speech and language impairments, mild motor developmental delay, wide variability in intellectual functioning, hypotonia, vision impairment, and behavioural problems such as attention/concentration deficits and hyperactivity^15,16^. Heterozygous pathogenic LoF variants in *SETBP1* have been independently identified by exome/genome sequencing in different cohorts of individuals with childhood apraxia of speech (CAS)^13,17^. In a comprehensive speech phenotyping study led by Morgan *et al.*^15^, poor communication was shown to be a central feature of *SETBP1* haploinsufficiency disorder. These findings further highlight the heterogeneity and complexity of SETBP1-related aetiologies.

*SETBP1* is expressed in numerous tissues including the brain, and multiple alternative transcripts encoding different isoforms have been found [OMIM 611060]. However, there is little existing knowledge concerning the expression and functions of the SETBP1 protein as well as its isoforms, with much of our understanding coming from studies of somatic variants in haematopoietic cells or overexpression systems. Accumulation of SETBP1 has been shown to reduce protease cleavage of its interactor oncoprotein SET, resulting in the formation of a SETBP1-SET-PP2A complex that inhibits PP2A phosphatase activity, thus promoting proliferation of leukaemic cells^6,7,18,19^. SETBP1 has chromatin remodeller functions, binding to AT-rich genomic regions via two AT-hooks, and has been shown to recruit a HCF1/KMT2A/PHF8 epigenetic complex in HEK cells overexpressing SETBP1^20^. Acting as a transcription factor, SETBP1 is able to activate or repress expression of genes such as *HOXA9, HOXA10, RUNX1, MYB* and *MECOM* in haematopoietic cells and in HEK cells overexpressing an SGS variant^20–23^. Nevertheless, the roles of SETBP1 in the developing brain and the pathophysiological pathways underlying germline pathogenic *SETBP1* variants remain elusive. Thus far, only three recent studies have investigated the molecular consequences of *SETBP1* disruptions in mouse and human neuronal models^8,20,24^, all of which described impairment in cell proliferation as a shared mechanism. Overexpression of human wild type SETBP1 and an SGS variant in mouse embryos via i*n utero* electroporation disrupted neuronal migration and neurogenesis in the neocortex^20^. In neural progenitor cells derived from patients with SGS, accumulation of SETBP1 promoted cell proliferation and induced DNA damage via SET stabilization that inhibited P53, and a PARP-1-dependent mechanism^8^. Investigations of neural progenitors in which *SETBP1* had been knocked out identified prolonged proliferation and distorted layer-specific neuronal differentiation with overall decrease in neurogenesis via the WNT/β-catenin signalling pathway^24^. However, these studies focused on a few SGS variants or *SETBP1* knockouts, leaving the majority of missense variants uncharacterized.

In 2017, Acuna-Hidalgo *et al.* identified four individuals carrying *SETBP1* variants in close proximity to the canonical degron [p.(Glu862Lys), p.(Ser867Arg), p.(Thr873Ile)] who showed a milder developmental phenotype with clinical characteristics that partially overlapped with classical SGS^6^. Additional individuals carrying missense variants in close proximity to the degron have since been reported in the medical literature. Nevertheless, the majority of the variants reported thus far are classified as variants of uncertain significance and their functional impacts have not been thoroughly characterized. Genotype-phenotype associations of germline *SETBP1* variants therefore remain unclear. The highly variable phenotypes and severity seen in these patients have made diagnoses difficult and precluded development of new, personalized therapies, often creating confusion among clinicians and patient families.

The current study addresses these important issues using a gene-driven approach. We recruited the largest cohort of individuals, to our knowledge, with a molecular diagnosis of *SETBP1* variants (missense variants and one in-frame deletion) outside the degron region (i.e. not affecting amino acids 868-871) who show clinical features that do not fit with the original SGS diagnostic criteria. We investigated the genotype-phenotype relationships of variants in the *SETBP1* gene by thorough phenotyping of clinical and speech/language features as well as functional follow-up of specific variants in cellular assays. We discovered that *SETBP1* variants outside the degron lead to a neurodevelopmental disorder characterised by a broad spectrum of clinical features, which are much milder than classical SGS despite partial overlaps, but in some cases more severe than *SETBP1* haploinsufficiency disorder. Using functional cellular assays, we showed that effects of SETBP1 variants involve a combination of mechanisms including deficits in protein degradation, ubiquitination and transcriptional control, independent of elevated protein levels. Overall, our findings point towards pathophysiological mechanisms that act via functional dosage of SETBP1 protein in a way that is dependent on variant location and specific change in amino-acid residues, rather than merely the amount of protein, explaining the clinical heterogeneity observed in patients.

## Material and Methods

### Molecular analyses

Peripheral blood samples were collected in a diagnostic context from the proband, and parents when available. Results originated from diagnostic or research-based genome sequencing, exome sequencing, gene panel testing for intellectual disability or epilepsy or via initial direct Sanger sequencing of the *SETBP1* gene based on clinical phenotypes, which in one case was followed by trio-based exome sequencing (details in Table S1).

### Patients and consent

This study analysed the medical data for patients with a variant in *SETBP1*. Only patients with missense variants outside the degron region (not affecting amino acids 868-871) of *SETBP1* were included in this study. The clinical data and patient materials were obtained through international collaborations involving clinicians from various countries (Table S1). Some of these collaborations were established via GeneMatcher^25^. Figure 2, Table 1, Table S1 and Figure S1 are not included in this preprint and are available from the corresponding author on request. Consent procedures and details of the IRB/oversight body that provided approval or exemption for the research described are described in the ethical statement.

**Table 1:** Summary of clinical phenotypes in individuals with *SETBP1* variants outside the degron. Not available in this preprint.

### Speech and language phenotyping

History of speech and language development was recorded. Speech was analysed for the presence of articulation, phonological disorder, stuttering, CAS^17,26^ and dysarthria^27,28^. Clinical diagnostic reports from local treating speech pathologists were used to validate the diagnoses with 100% agreement. The Intelligibility in Context Scale^29^ examined how often the participant’s speech was understood, with a 5-point scale of responses ranging from never to always. Children were assessed for verbal, written and social language with the standardised Vineland Adaptive Behaviour Scales-Parent/Caregiver III^30^. Adult participants were assessed for receptive language with the Peabody Picture Vocabulary Test-4^31^ and clinical speech pathology (ST, RB, AM) and neurology (IS) assessment determined expressive language, social skills and written language ability. Table 2 is not included in this preprint and is available from the corresponding authors on request.

**Table 2:** Summary of speech phenotypes in individuals with *SETBP1* missense variants outside the degron. Not available in this preprint.

### Spatial clustering analysis of missense variants

All independently observed *SETBP1* missense variants outside the canonical degron were included in the spatial clustering analysis^32,33^. The geometric mean was computed over the locations of observed missense variants in the canonical transcript of *SETBP1* (9,899 bp; NM_015559.2) and subsequently compared to 100,000,000 permutations by randomly redistributing the variant locations over the total size of the *SETBP1* coding region and calculating the resulting geometric mean from each of these permutations. The corrected *p*-value was computed by checking how often the observed geometric mean distance was smaller than the permutated geometric mean distance and considered significant if <0.05.

### Cell culture and transfection

HEK293T/17 cells (CRL-11268, ATCC) were cultured in Dulbecco’s modified Eagle’s medium (DMEM) (Gibco) supplemented with 10% foetal bovine serum (FBS) (Gibco) and 100 U/ml Penicillin-Streptomycin (Thermo-Fisher) at 37°C and 5% CO2. For immunofluorescence analysis, cells were seeded onto coverslips coated with 100 μg/ml poly-L-lysine (Merck-Millipore). Fibroblast cell lines were established from skin biopsies of patients with *SETBP1* variants and controls at the Cell Culture Facility, Department of Human Genetics at Radboud university medical center. Human dermal fibroblasts were cultured in DMEM supplemented with 20% FBS, 100 U/ml Penicillin-Streptomycin and 1% sodium pyruvate (Thermo-Fisher) at 37°C and 5% CO2. Transfections were performed using transfection reagent GeneJuice (Merck-Millipore) following the manufacturer’s instructions or polyethyenimine (PEI) (Sigma) in a 3:1 ratio with the total mass of DNA transfected.

### DNA Constructs and site-directed mutagenesis

The full-length SETBP1 construct fused to a C-terminal Myc-DDK tag under a human CMV promoter (pCMV-Entry-SETBP1) was purchased from Origene (RC229443). Constructs carrying *SETBP1* variants (pCMV-Entry-SETBP1) were generated using QuikChange Lightning Site-Directed Mutagenesis Kit (Agilent) following the manufacturer’s protocol. To generate constructs carrying *SETBP1* fused to an YFP-tag (pYFP-SETBP1), *SETBP1* cDNAs were first subcloned using EcoRI/XhoI restriction sites into a modified pEGFP-C2 vector (Clontech) where the N-terminal EGFP-tag was then replaced with an YFP-tag using BshTI/Bsp1407I restriction sites. To generate AT-rich reporter vectors used in luciferase reporter assay, 100 µM sense and antisense single-stranded oligonucleotides carrying 5’-AAAATAA-3’ or 5’-AAAATAT-3’ repeats were first annealed using 2x annealing buffer [20mM Tris (Thermo-Fisher), 2mM EDTA (Sigma), 100mM NaCl (Sigma), pH 8.0]. Annealed oligonucleotides were phosphorylated using 1x T4 DNA ligase buffer and 30 units of T4 PNK (New England Biolabs), then inserted into a pGL4-luc2 vector using KpnI/HindIII restriction sites. A firefly luciferase reporter construct containing FOXP2 promoters, TSS1 and TSS2, respectively were a gift from the Vernes group^34^. For mammalian one-hybrid (M1H) assays, a pGL4-luc2-GAL4UAS-adenovirus promotor reporter construct and a pGL4.23 construct with an adenovirus major late promotor (pBIND2) were generated as previously described^35^. *SETBP1* cDNAs were subcloned from pCMV-Entry-SETBP1 into the empty pBIND2 using SaII/NotI restriction sites generating pBIND2-SETBP1 expression vectors. A GFP-SET construct was generated as previously described^6^. All constructs were verified by Sanger sequencing. Primer sequences are listed in Table S11.

### Immunofluorescence microscopy

HEK293T/17 cells grown on poly-L-lysine-coated coverslips were transiently transfected with 500ng of a pCMV-SETBP1 or pYFP-SETBP1 construct. Cells were fixated 48 hours after transfection using 4% paraformaldehyde solution (Electron Microscopy Supplies Ltd) for 20 minutes at room temperature (RT). Human dermal fibroblasts were grown on coverslips coated with 1 µg/ml fibronectin (Corning) and fixated as above. Fixated cells were permeabilized with 0.4% Triton-X-100/PBS for 20 minutes at RT. Following incubation in a blocking solution for 1 hour at RT, cells were incubated in primary antibodies overnight at 4°C and then with conjugated secondary anti-IgG antibodies for one hour at RT. Hoechst 33342 (Invitrogen) was used for nuclear staining, before mounting with VECTASHIELD® Antifade Mounting Medium (Vectorlab). Fluorescence images were obtained using an LSM880 AxioObserved confocal microscope (Zeiss). For images of single nuclei, the Airyscan unit (Zeiss) was used with a 4.0 zoom factor. Images were analyzed with an ImageJ “Analyze particle” plugin. A list of antibodies used can be found in Table S12.

### Co-immunoprecipitation

HEK293T/17 cells grown in a 10-cm culture dish were transiently transfected with constructs expressing FLAG-SETBP1 only or together with GFP-SET under a human CMV promoter. An empty pCR2.1-TOPO (Invitrogen) was used as a filler to top up to 12 µg DNA in total per transfection. Cells were lysed in 1ml of Pierce® IP Lysis Buffer (Thermo-Fisher) supplemented with protease inhibitors (Roche) and 1% PMSF (Thermo-Fisher) 48 hours post-transfection. All following steps were performed at 4°C. Cell lysates were incubated for 10 minutes under rotation, centrifuged at 13200 rpm for 20 minutes. The supernatant was then quantified with a Pierce BCA protein assay kit (Thermo-Fisher). 5% protein lysate was collected as input and denatured in Laemmli buffer (Bio-Rad). 400 µg protein was immunoprecipitated with 30 µl FLAG-agarose beads (Thermo-Fisher) or 25 µl GFP-Trap (Chromotek) on a rotating wheel overnight. Beads were then washed three times with PBS and eluted with Laemmli buffer for immunoblotting analysis.

### Immunoblotting and band intensity quantification

HEK293T/17 cells grown in a 6-well culture plate were transiently transfected with 3 µg DNA. Cells were lysed in 1 ml of Pierce® RIPA Buffer (Thermo-Fisher) supplemented with protease inhibitors (Roche) and 1% PMSF (Thermo-Fisher) 48 hours post-transfection. Total protein was quantified using a BCA assay. Proteins were resolved on 4 –15% Tris-Glycine gels and transferred to PDVF membranes (Bio-Rad). After blotting, membranes were incubated overnight at 4°C with the appropriate primary antibodies, followed by HRP-conjugated secondary antibodies (Table S12). Proteins were visualized using the Novex ECL Chemiluminescent Substrate Reagent kit (Invitrogen) or SuperSignal West Femto Maximum Sensitivity Substrate (Thermo-Fisher) and the ChemiDoc XRS+ System (Bio-Rad). Band intensity was quantified using ImageJ. Background-subtracted band intensity was divided by the background-subtracted band intensity of β-actin for normalization. For quantification of (co-) immunoprecipitation experiments, the background-subtracted band intensity of the immunoprecipitated fraction was normalised with respect to the input fraction for each condition.

### Fluorescence-based quantification of protein stability and degradation

HEK293/T17 cells were transfected in triplicate in clear-bottomed black 96-well plates with YFP-tagged SETBP1 variants. After 24 hours, cycloheximide (Sigma) at a final concentration of 50 μg/ml was added, MG132 (R&D Systems) at 5 µg/ml and Bafilomycin A1 (R&D Systems) at 100 nM. Cells were incubated at 37°C with 5% CO2 in the Infinite M200Pro microplate reader (Tecan), and the fluorescence intensity of YFP (Ex: 505 nm, Em: 545 nm) was measured over 24 hours at 3-hour intervals.

### Luciferase reporter assay and mammalian-one-hybrid (M1H) assay

HEK293/T17 cells were seeded in clear-bottomed white 96-well plates (Greiner Bio-One) and transfected in triplicates using GeneJuice (Merck-Millipore). Cells were co-transfected with 350 ng of firefly luciferase reporter construct containing six repeats of AT-rich consensus sequence (5’-AAAATAA-3’ or 5’-AAAATAT-3’) or *FOXP2* promoters (TSS1/TSS2)^34^, 6.5ng of pGL4.74 Renilla luciferase normalization control, and 700 ng of pYFP construct with or without SETBP1. For M1H assay, cells were co-transfected with 1,433 ng (50 nM) of expression construct containing only GAL4 or GAL4 including SETBP1, 416 ng of reporter construct with or without GAL4-binding site, and 165 ng of Renilla luciferase normalization control. After 48 hours, firefly luciferase and Renilla luciferase activities were measured using a Dual-Luciferase Reporter Assay system (Promega) according to manufacturer’s instructions at an Infinite M Plex Microplate reader (Tecan).

### Cell proliferation assay

Fibroblasts were seeded in triplicate in clear-bottomed black 96-well plates (Greiner Bio-One). Cell proliferation was measured using a CyQUANT™ Direct Cell Proliferation Assay (Thermo-Fisher) according to manufacturer’s instructions at an Infinite M Plex Microplate reader. Fluorescence intensity were measured daily for four days. Background fluorescence was subtracted and values were normalized to day 0. Growth curves were plotted as fluorescence versus time.

### RNA sequencing (RNA-seq) and data analysis

Three technical replicates of each fibroblast cell line were included in the RNA-seq experiment. Total RNA was extracted from one to two million cells. 1 μg of total RNA extracted (Qiagen) was used to generate RNA libraries using NEBNext® UltraTM RNA Library Prep Kit for Illumina® (New England Biolabs) following the manufacturer’s protocol and index codes were added to attribute sequences to each sample. The libraries were sequenced on a NovaSeq6000 platform using 150bp pair-end reads. Image processing and basecall was performed using the Illumina Real Time Analysis Software. Fastq files were aligned to the human genome (GRCh38/hg39, Ensembl) using HISAT2^36^ software together with the corresponding splice junctions Ensembl GTF annotation. At least 89% of the reads for each sample were uniquely mapped to the human genome entailing at least 36.4 million unique reads in the sample with the lowest sequencing depth. Principal Component Analysis (PCA) was performed using the SeqMonk tool (version 1.47.1). Differential gene expression was performed using DESeq2^37^ and intensity differences [*p*>0.05, false discovery rate (FDR)] were calculated via the SeqMonk tool (version 1.47.1) for female and male samples (patients versus controls) separately. Gene set enrichment analysis of the differentially expressed genes (DEGs) that were present in both female and male samples was performed using GSEA software (version 4.1.0)^38^ using gene sets for biological pathways (c5.go.bp.v7.2.symbols.gmt) with 1000 permutations and multiple testing correction (*p*>0.05, FDR). Gene ontology analysis was performed using g:GOSt within the g:Profiler web server^39^. A list of expressed genes in our fibroblast samples was used as the custom background for gene ontology analysis with multiple testing correction (*p*<0.05, Benjamini-Hochberg FDR). Degrees of overlap between the differentially expressed genes in our fibroblast samples and autism (v.0.22)- or intellectual disability (ID, v.3.2)-associated genes in the PanelApp database were calculated using a Fisher’s exact test (*p*>0.05). A list of expressed genes in our fibroblast samples was used as the custom background. Datasets generated and analysed in the current study are not included in this preprint, and are available from the corresponding authors on request.

### Quantitative real-time PCR

Total RNA was extracted with Qiazol or an RNeasy Plus mini kit (Qiagen) following manufacturer’s protocols. 1 µg of total RNA was used to synthesis cDNA using SuperScriptIII Reverse Transcriptase (Thermo-Fisher). Real-time PCR was performed using iQ SYBR Green Supermix (Bio-Rad) at a CFX384 Touch Real-Time PCR Detection System (Bio-Rad).

### Statistical analysis of cell-based functional assays

Statistical analyses for cell-based functional assays were done using a one- or two-way ANOVA followed by a Bonferroni, Tukey, or Dunnett *post-hoc* test, with GraphPad Prism (version 8.0) Software.

## Results

### *SETBP1* variants outside the canonical degron of the SETBP1 protein cluster in the SKI domain

In our study, we identified 18 unrelated individuals carrying rare heterozygous variants with uncertain functional impact in *SETBP1* (NM_015559.2), a gene under constraint against loss-of-function and missense variation [pLoF: o/e = 0.02 (0.01 – 0.11); missense: o/e = 0.9 (0.84 – 0.95); gnomAD v.2.1.1^40^] (Figure 1A; Tables 1, S1 and S3). Variants were identified via exome or genome sequencing. In one case, the variant was first identified with direct Sanger sequencing of the *SETBP1* gene based on clinical observations in the patient followed by trio-based exome sequencing. None of the 18 individuals met the diagnostic criteria of Schinzel-Giedion Syndrome (SGS)^41^. Fifteen individuals carried a *de novo SETBP1* variant, while the other two had inherited a variant from an affected parent. One individual harboured a missense variant that was not inherited from the mother; the father was unavailable for testing. Among the 15 individuals with a *de novo* variant, 14 carried a missense variant and one had an in-frame deletion [c.2885_2887del(CCA) p.(Thr962del)]. Within our cohort, there were multiple cases of recurrent identical *de novo* variants including c.2572G>A p.(Glu858Lys) found in four children and c.2584G>A p.(Glu862Lys) in two individuals, revealing an independent mutational hotspot located in close proximity to the canonical degron region. None of the *SETBP1* variants included in our study were present in the gnomAD v.2.1.1 database. Two individuals also carried variants affecting other known disease genes. In proband 3, who has a *de novo* c.2561C>T p.(Ser854Phe) *SETBP1* variant, an *EHMT1* variant was identified. In proband 1, who has a c.1332C>G p.(Ser444Arg) *SETBP1* variant, an inherited 5q22.31 dup and *WWOX* variants were identified. However, these variants in probands 1 and 3 are unlikely to be pathogenic based on the patients’ clinical features, the inheritance model and the lack of functional impact observed in assays performed in cellular models (unpublished data with the *EHMT1* and *WWOX* variants).

**Figure 1:**
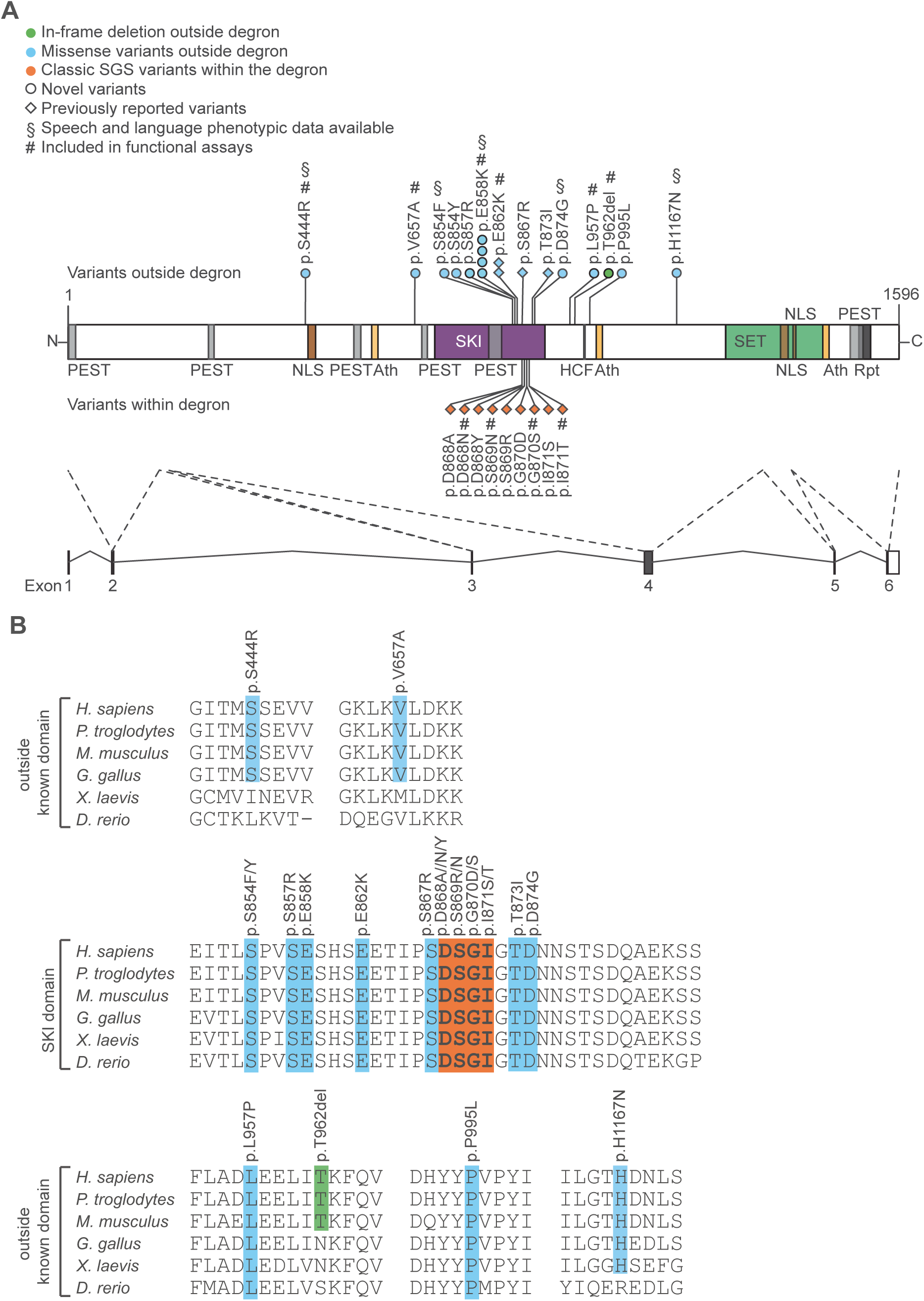
*SETBP1* variants outside the degron cluster in the SKI domain and are highly conserved across species. (A) Schematic representation of the SETBP1 protein (UniProt: Q9Y6X0) indicating the locations of variants included in this study. These comprise eleven novel germline variants (circles) including ten missense variants (blue) and one in-frame deletion (*de novo*, green) outside the canonical degron. Three missense variants outside (blue) and nine within (orange) the degron that were previously reported (diamond) are also annotated in the schematic. § represent variants for which speech phenotyping data were available. # represent variants included in functional assays. Five exons (black bars) encode isoform A of the protein (NP_056374.2, 1596 amino acids). The SETBP1 protein sequence contains three AT hook domains (Ath; orange; amino acids 584–596, 1016–1028, 1451–1463), a SKI homologous region (SKI; purple; amino acids 706–917), an HCF1-binding motif (HCF; black; amino acids 991–994), a SET-binding domain (SET; green; amino acids 1292–1488), three bipartite NLS motifs (brown; amino acids 462-477, 1370-1384, 1383-1399), six PEST sequences (grey; amino acids 1-13, 269-280, 548-561, 678-689, 806-830, 1502-1526) and a repeat domain (Rpt; black; amino acids 1520–1543)^7,19,20^. An overview with variant details per subject is provided in Table 1. (B) Sequence alignment of the region containing part of the SETBP1 amino acid sequence in human (Uniprot: Q9Y6X0), chimpanzee (H2QEG8), mouse (Q9Z180), chicken (A0A1D5PT15), African clawed frog (F6TBV9) and zebrafish (B0R147). The canonical degron is highlighted in bold. Residues in which germline missense variants located within (orange), outside the degron (blue) and deleted by an in-frame deletion (green) are highlighted.

**Figure 2:** Clinical evaluation of individuals with *SETBP1* variants. Not available in this preprint.

### *SETBP1* variants outside the canonical degron cause a broad spectrum of clinical features

An overview of the main clinical features per individual is shown in Table 1. More detailed data for each patient are shown in Table S1. Individuals (n=5) with a variant in close proximity to the degron within the SKI domain (affecting amino acids 862-874, excluding the degron 868-871) showed severe or profound intellectual disability and severe motor impairment with inability to walk without support. Four of these individuals were unable to speak. Three individuals showed spasticity. Focal and tonic-clonic seizures were noted in two of these individuals. Two individuals showed renal abnormalities: one had mild kidney dilatation, another medullary cystic kidneys. Shared facial features were present in at least three out of the five individuals, including prominent ears, shallow orbits, midface retraction and microcephaly. Overall, the phenotypes of these five children did not fulfil the original Lehman et al. criteria for SGS^41^. However, based on the severity of the phenotypes and facial features, they appeared more similar to (but less severe than) SGS compared to *SETBP1* haploinsufficiency disorder.

Individuals (n=7) with variants (amino acids 854-858) located slightly further away from the degron but still within the SKI domain showed mild or moderate intellectual disability. Two out of four individuals with a c.2572G>A p.(Glu858Lys) variant were minimally verbal (see the speech and language section; the relevant data on this from the other two individuals were unavailable). All seven individuals showed motor delay, but the degree was much milder compared to the aforementioned cases. All those aged four years and older were able to walk without support, although one individual walked only limited distances. One individual of 3.5 years did not walk yet. Absence seizures were noted in two of these cases. One individual had asplenia. One individual had a non-progressive heart tumour of unknown origin. These individuals did not show recognizable or similar facial features, nor did they show overlapping facial features with either classical SGS or *SETBP1* haploinsufficiency disorder (data not shown).

The individuals with inherited variants located furthest away from the degron and the SKI domain [c.1332C>G p.(Ser444Arg) and c.1970T>C p.(Val657Ala)] showed mild intellectual disability or a low non-verbal IQ. In both cases, parents carrying the variant were similarly affected. For the remainder of the study, we therefore did not distinguish these inherited variants from the *de novo* variants outside the degron. All variants outside the degron were considered in functional assays as one group versus those within the degron (causing classical SGS).

Variants located outside the SKI domain (n=4) were associated with a more variable clinical phenotype. One individual with an in-frame deletion removing a threonine residue c.2885_2887del(CCA) p.(Thr962del) showed a severe phenotype with severe speech delay, inability to walk and tonic-clonic seizures. This individual had bilateral ptosis and had surgery to the right upper eyelid and left strabismus surgery. This proband’s facial features appeared similar to the individual with the c.2984C>T p.(Leu957Pro) variant. They both showed a round face, blepharophimosis, hypertelorism and a short nose with a bulbous tip, features also often noted in individuals with *SETBP1* haploinsufficiency disorder. The latter individual had a less severe neurodevelopmental phenotype. This individual started to walk at 22 months and was able to use sign language at the age of three years.

### Individuals carrying SETBP1 variants outside the canonical degron show generally low speech/language ability

Speech/language data were available from seven individuals in this cohort, four of whom carry a variant within the SKI domain close to the degron (probands 3, 6, 8 and 14) while three individuals harbour a variantlocated far from the degron and outside the SKI domain (probands 1 and 18, and affected mother of proband 1). In this cohort, speech development during infancy was characterised by limited babbling. A history of early feeding difficulties was also present for two children (probands 3 and 18). Language ability was generally low across the group (n=7) for all subdomains including expressive, receptive, written and social language (Table 2 and Figure 3). The youngest children (< 9 years of age; probands 3, 6, 8 and 18) present with a severe speech and language impairment. They are minimally verbal, defined as the presence of less than 50 spoken words (Table 2), and augment verbal communication with sign language, gestural communication and digital devices. The speech motor system is impaired across all individuals in the group, with Childhood Apraxia of Speech (CAS) the most common diagnosis (n=5), followed by mild dysarthria (proband 1 and affected mother) (Table 2). CAS features included hesitancy, groping, inconsistency of production, increased errors with increasing word length, simplified word and syllable structures relative to age, and vowel and prosodic errors. Dysarthria was typically characterised by slower rate of speech, imprecision of consonants, altered nasal resonance and monotonous speech. The adult participants (proband 1 and affected mother) had a history of poor speech development but are now able to hold appropriate conversations and speak in full sentences with speech that is usually to always intelligible. All individuals in this cohort who performed speech/language assessment are attending (probands 3, 6, 8 and 18), or had attended (probands 1 and 14, and affected mother of proband 1), speech therapy.

**Figure 3:**
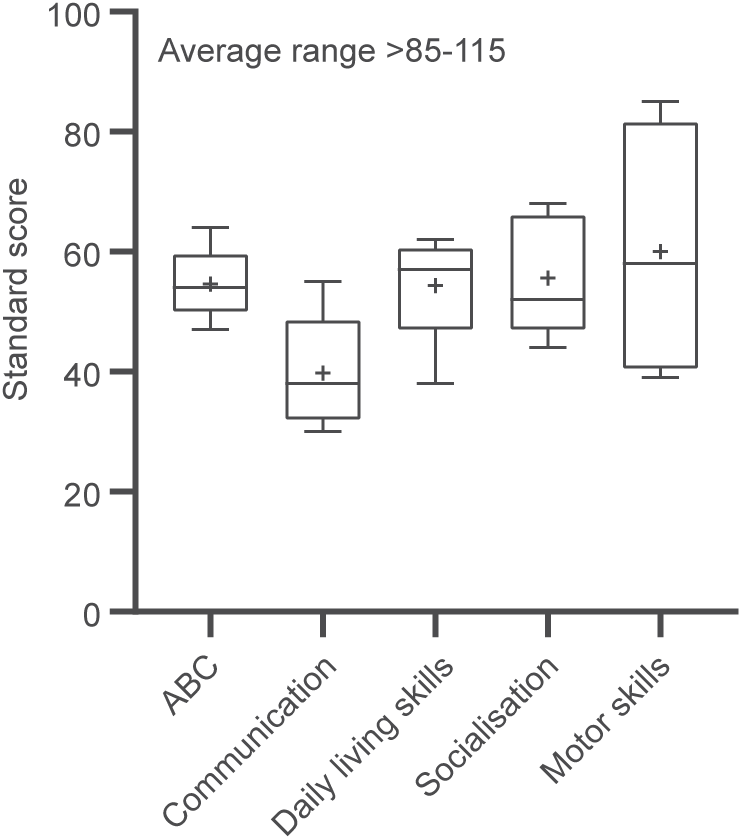
Performance on Vineland-3 subtests. Lines denote median scores; X denotes mean scores; ABC, Adaptive Behaviour Composite. Standard scores between 85 and 115 are considered within the average range, with a mean of 100 and a standard deviation of 15.

### *SETBP1* variants outside the canonical degron are predicted to be damaging and cluster in the SKI domain

We used an array of computational tools to predict the functional effects of the observed *SETBP1* variants. Among the 14 variants observed in 18 individuals, eight were located in the SKI domain while six were outside any known functional domain (Figure 1A). Using a spatial clustering analysis^33^, we showed that these variants outside the degron significantly clustered in exon 4 of the canonical *SETBP1* transcript (NM_015559.2) corresponding to the SKI domain (corrected *p*-value = 9.99e-09, Bonferroni correction). All of the mutated amino acid sites were highly conserved across species with the exception of the threonine residue at position 962, which was conserved only in mammals (Figure 1B). All observed variants were predicted to be (likely) pathogenic by PolyPhen-2 and/or SIFT, and showed CADD-PHRED scores above 21 (Table S2).

We went on to use the MetaDome web tool (v.1.0.1) to visualise all *SETBP1* missense variants in the tolerance landscape of the gene (Figure S2A and B). Variants in the SKI domain are located in the regions of high intolerance while the remaining variants, including those adjacent to the HCF1-binding site, are located in the less tolerant regions (Figure S2A and B). Of note, *SETBP1* does not have a particularly high Z-score (1.1) for missense variants in the gnomAD database (v.2.1.1), indicating that the complete coding region of this gene is not extremely intolerant to missense variation. This observation is consistent with the results from the MetaDome analysis, which show that only a few regions of SETBP1 have a high intolerance for missense variants, including the part of the SKI domain in which the majority of the observed variants are located (Figure S2B). We observed that 33% of the amino acid residues that were mutated by a germline *SETBP1* missense variant were also mutated in somatic cells, particularly in haematopoietic and lymphoid cells, according to the COSMIC database (Catalogue of Somatic Mutations in Cancer v.94), including the recurrent missense variant p.(Glu858Lys). While missense variants within the degron (affecting amino acids 868-871) showed different frequencies in germline and somatic cells, consistent with the previously reported higher functional threshold in somatic cells^6^, all observed missense variants outside the degron showed similar frequencies in germline and somatic cells (Figure S3 and Table S3).

### *SETBP1* missense variants outside the degron cause variable disruption of protein degradation in contrast to accumulating stable SGS variants

We went on to study the functional consequences of a representative selection of variants across *SETBP1* on protein localization, protein stability and degradation, cell proliferation and transcriptional activity using HEK293T/17 cells transiently transfected with *SETBP1* expression constructs, as well as in patient fibroblasts. Based on the location and their distance to the canonical degron (Figure 1A), we included in our assays two missense variants located furthest from the SKI domain [p.(Ser444Arg) and p.(Val657Ala)], three missense variants close to the degron [p.(Glu858Lys), p.(Glu862Lys)], and p.(Leu957Pro)], and one *de novo* in-frame deletion [p.(Thr962del)]. In addition, we included in our assays four classical SGS variants located within the canonical degron [p.(Asp868Asn), p.(Ser869Asn), p.(Gly870Ser), and p.(Ile871Thr)] for functional comparisons to those outside the degron.

We first assessed the abundance of FLAG-tagged SETBP1 in transfected HEK293T/17 cells, comparing cells with variant expression constructs to those with a wild type construct. All variants showed higher SETBP1 protein levels than the wild type (Figures 4A, 4B and S4A). Next, we hypothesized that missense variants outside the degron might alter SETBP1 stability. We therefore treated HEK239T/17 cells expressing YFP-tagged SETBP1 with cycloheximide to inhibit translation and measured relative fluorescence intensity over 24 hours. We found that all classical SGS variants (within the degron) showed increased protein stability whereas all variants outside the degron displayed similar stability to wild type (Figures 4C and S6). To evaluate the impact of variants on proteasome-mediated degradation, we treated HEK293T/17 cells expressing YFP-tagged SETBP1 with proteasome inhibitor MG132. Surprisingly, the classical SGS variants did not show impaired proteasome degradation, except for p.(Gly870Ser) (Figures 4C and S7), unlike previously reported^6^. Moreover, three variants outside the degron [p.(Ser444Arg), p.(Glu858Lys), and p.(Leu957Pro)] demonstrated disrupted proteasome degradation to various extents (Figure 4C). To assess whether the degradation of *SETBP1* variants might be compensated by other protein degradation pathways, such as mTOR-dependent autophagy, we used the autophagy inhibitor BafilomycinA1 to treat HEK293T/17 cells expressing YFP-tagged SETBP1. While the majority of variants outside the degron [p.(Ser444Arg), p.(Val657Ala), p.(Glu858Lys), and p.Leu957Pro)] differed significantly in degradation via autophagy, most of the SGS variants were similar to wild type (Figures 4C and S8). It is noteworthy that the direction and extent of degradation of the variant proteins was variable and appeared to depend on the distance of the variant from the degron region. Consistent with the overexpression system, patient fibroblasts [p.(Glu858Lys), p.Leu957Pro, and p.(Thr962del)] also showed significantly more abundant SETBP1 protein compared to healthy controls, although mRNA expression appeared to be more variable (Figure 4D). Interestingly, protein stability and degradation of the in-frame deletion p.(Thr962del) was not affected (Figures 4C and S6A), suggesting that it might operate via a different pathophysiological mechanism, in spite of increased abundance.

**Figure 4:**
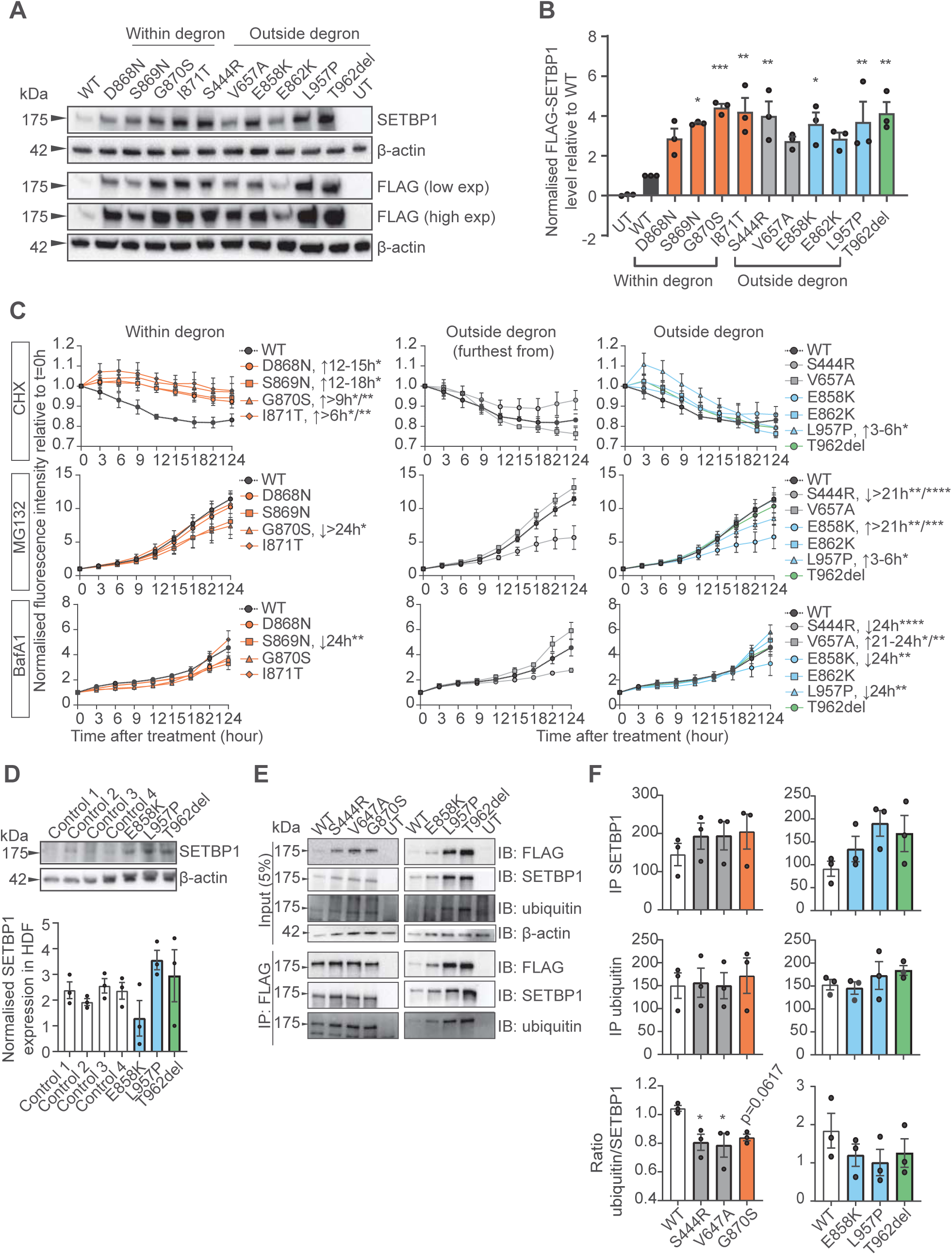
Variable impairment of SETBP1 degradation via proteasome and autophagy pathways is associated with partial reduction of ubiquitination. (A) Immunoblot of whole cell lysates of HEK293T/17 cells expressing FLAG-tagged SETBP1 variants probed with anti-SETBP1 and anti-FLAG antibodies. β-actin was used as a loading control. Low and high exposures of immunoblot probed with anti-FLAG antibody are shown. Representative blots of three independent experiments are shown. (B) Quantification of protein levels of FLAG-tagged SETBP1 variants normalised to β-actin. Values are expressed relative to wild type (WT) and represent the mean ± SEM of three independent experiments (\**p*< 0.05, \*\**p*< 0.01, \*\*\**p*< 0.001, using one-way ANOVA and a *post-hoc* Dunnett’s test). (C) Relative fluorescence intensity of YFP-tagged SETBP1 variants overexpressed in HEK293T/17 cells treated with translation inhibitor cycloheximide (CHX; 50µg/ml; top), proteasomal degradation inhibitor MG132 (5µg/ml; middle), or autophagy inhibitor Bafilomycin A1 (BafA1; 100nM; bottom). An equal volume of DMSO was used as a vehicle control. Fluorescence intensity was measured for 24 hours with three-hour intervals and normalised to an mCherry transfection control. Values are expressed relative to t= 0 hour and represent the mean ± SEM of three independent experiments, each preformed in triplicate (\**p*< 0.05, \*\**p*< 0.01, \*\*\**p*< 0.001, **** *p*< 0.0001; two-way ANOVA and a *post-hoc* Dunnett’s test). (D) Immunoblot of whole cell lysates of control and patient human dermal fibroblasts (HDF) probed with anti-SETBP1 antibody. β-actin was used as a loading control (top). Normalised *SETBP1* transcript expression (bottom) in control and patient HDFs. Bars represent mean ±SEM of three independent experiments (n.s., \**p*<0.05, versus WT; one-way ANOVA and a *post-hoc* Dunnett’s test). (E) Immunoprecipitation of FLAG-tagged SETBP1 wild type and variants using FLAG-conjugated magnetic agarose and blotted with an anti-FLAG, anti-SETBP1 or anti-ubiquitin antibody. β-actin was used as a loading control. (F) Quantification of SETBP1 (top), ubiquitin (middle) in FLAG-IP fractions for inherited (outside degron, grey) and SGS variants (orange, left), and *de novo* variants outside the degron (right). Ratio of ubiquitin/SETBP1 in the FLAG-IP fractions was plotted (bottom). Bars represent mean ±SEM of three independent experiments (\**p*<0.05, versus WT; one-way ANOVA and a *post-hoc* Dunnett’s test).

*In silico* modelling of germline variants within the canonical degron that cause classical SGS has suggested effects on the interaction between the degron of β-catenin, which has similar sequence to the βTrCP1 binding site in the SETBP1 degron, and ubiquitin E3 ligase βTrCP1^6^. To investigate whether the observed differences in protein degradation were due to alterations in SETBP1 ubiquitination, we performed immunoprecipitation of FLAG-SETBP1 and assessed its ubiquitin level. Even though impaired proteasome degradation and autophagy were observed in two classical SGS variants and several variants outside the degron (Figure 4C), ubiquitination was not significantly reduced in the majority of the tested variants [p.(Gly870Ser), p.(Glu858Lys), p.(Leu957Pro), and p.(Thr962del)] (Figures 4E and 4F). Intriguingly, variants furthest from the degron showed significantly lower SETBP1 ubiquitination compared to wild type (Figures 4E and 4F), consistent with the degradation assay results (Figure 4C). Although based on *in silico* modelling of interaction with ubiquitin E3 ligase βTrCP1^6^, the classical SGS variant p.(Asp868Asn) would be expected to show the strongest disruption in SETBP1 degradation, followed by p.(Gly870Ser), we did not see such a pattern in our proteasome and autophagy inhibition assays, nor in the level of ubiquitination. Taken together, these results suggest that accumulations of SETBP1 protein observed for a subset of variants are caused by variable disruptions in SETBP1 protein degradation via the proteasome and autophagy pathways. Other mechanisms are likely to contribute to higher SETBP1 protein levels in addition to pathways involving ubiquitination.

### *SETBP1* variants outside the degron, but not SGS variants, reduce binding to AT-rich DNA sequences and transcriptional activation

SETBP1 can bind to genomic DNA via its AT-hooks and function as a regulator of transcription^20^. We went on to assess the effects of SETBP1 variants on the capacity of the protein to bind to AT-rich DNA sequences, using a luciferase reporter system. We generated two luciferase reporters inserted respectively with six repeats of the previously reported consensus AT-rich DNA binding sequences of SETBP1 (5’-AAAATAA-3’ or 5’-AAAATAT-3’)^20^. The majority of the variants tested could still bind to AT-rich DNA sequences (Figure 5A). Of note, both variants [p.(Leu957Pro) and p.(Thr962del)] located close to the HCF binding domain (amino acids 991-994) showed significantly reduced AT-rich sequence binding capacity (Figure 5A).

**Figure 5:**
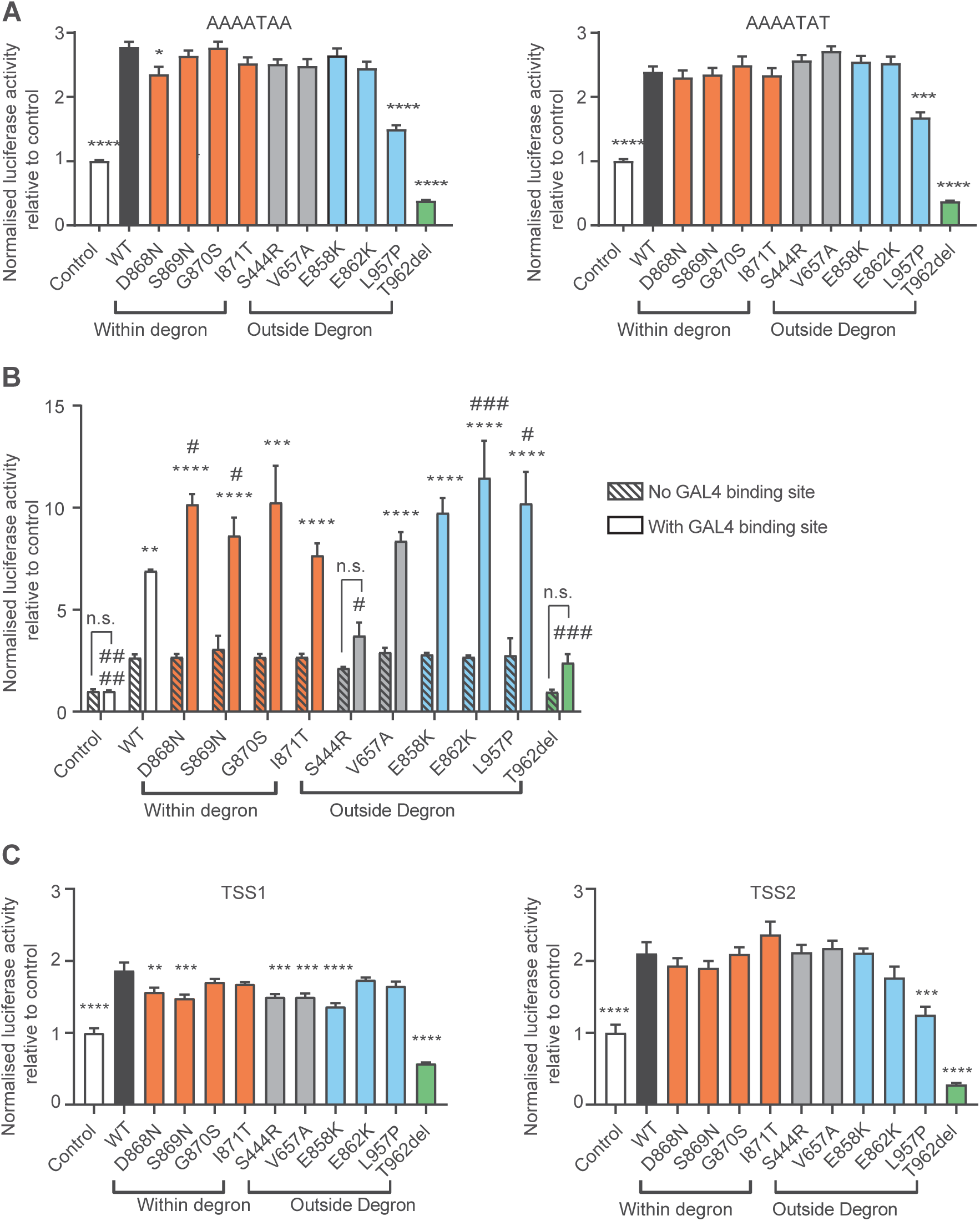
Genotype-specific reduction of binding capacity to AT-rich DNA sequences and transcriptional activation for *SETBP1* variants outside the degron. (A) Results of luciferase assays with constructs containing WT and *SETBP1* variants, and the reporter constructs with the consensus SETBP1 binding sequences. Values are expressed relative to the control condition which used a pCMV-YFP construct without SETBP1. (B) Results of the M1H assay for SETBP1 transcriptional regulatory activity with WT and *SETBP1* variants fused with an N-terminal GAL4 in combination with a reporter construct with or without the GAL4-binding site. Values are expressed relative to the control condition which used a pBIND2-GAL4 construct without SETBP1. (C) Results of luciferase assays with constructs containing WT and *SETBP1* variants, and reporter constructs with *FOXP2* promoters: TSS1 (left) and TSS2 (right). Values are expressed relative to the control condition which used a pCMV-YFP construct without SETBP1. All graphs for luciferase assays show the mean ± SEM of three independent experiments, each performed in triplicate (\**p*<0.05, \*\**p*<0.01, \*\*\**p*<0.001, \*\*\*\**p*<0.0001 versus WT; one-way ANOVA and a *post-hoc* Dunnett’s test). All graphs for the mammalian-one-hybrid assay show the mean ± SEM of three independent experiments, each performed in triplicate (\**p*<0.05, \*\*\**p*<0.001, \*\*\*\**p*<0.0001 versus reporter without GAL4-binding site; ^#^*p*<0.05, ^###^*p*<0.001, ^####^*p*<0.0001 versus WT; two-way ANOVA and a *post-hoc* Tukey’s test).

We next used a mammalian-one-hybrid (M1H) assay to further delineate whether SETBP1 can induce transcriptional activity in the proximity of promoter regions without direct DNA binding. Wild type SETBP1 fused with GAL4 showed significantly increased luciferase activity compared to empty controls and a reporter construct without a GAL4-binding site (Figure 5B). These results confirmed the capacity of the protein to activate transcription in the vicinity of a promoter region without direct binding to DNA, consistent with its role as a chromatin remodeller. The majority of the variants could activate transcription (Figure 5B). Interestingly, two SGS variants and two variants close to the degron [p.(Glu862Lys) and p.(Leu957Pro)] showed significantly higher transcriptional activity compared to wild type (Figure 5B). In contrast, the two variants furthest from the degron [p.(Ser444Arg) and p.(Thr962del)] failed to activate transcription, appearing to be loss-of-function (Figure 5B).

Moreover, our experiments identified *FOXP2,* rare genetic disruptions of which lead to CAS^42–44^, as a novel direct transcriptional target of SETBP1 (Figure 5C). Most of the variants that we tested led to reduced *FOXP2* transcription activation (Figure 5C). Notably, p.(Thr962del), which lacks only one threonine residue in the encoded SETBP1 protein, resulted in complete loss of function in all of our luciferase reporter assays, highlighting the importance of this residue for SETBP1 transcriptional activity. SETBP1 has been shown to be a largely nuclear protein and so its potential mislocalization could lead to disruption of its function. However, we found that all *SETBP1* variants localized to the nucleus as puncta similar to wild type when assessed in transiently transfected HEK293T/17 cells and endogenously in patient fibroblasts (Figure S5). Taken together, these data suggest that pathogenic *SETBP1* variants outside the degron reduce AT-rich DNA binding capacity and transcriptional activity of SETBP1 within the nucleus while preserving gross intracellular localization.

### Patient fibroblasts carrying *SETBP1* variants outside the degron show increased proliferation

Increased cell proliferation has been reported in EBV-transformed lymphoblastoid cell lines (LCLs) derived from patients carrying germline classical SGS variants^6^ and in leukaemic cells with somatic *SETBP1* variants that drive development of myeloid malignancies^6,45^. We therefore investigated the proliferation of fibroblasts derived from three individuals carrying a germline *SETBP1* variant outside the degron [p.(Glu858Lys), p.(Leu957Pro), and p.(Thr962del)] compared to those from sex-matched healthy controls. For two of the three variants, we observed that fibroblasts displayed significantly faster proliferation (Figures 6A and S10A) and shorter doubling time than healthy controls in a time course experiment (Figure 6B). Interaction between SETBP1 and SET has been shown to stabilise SET, protecting SET from cleavage by protease, subsequently inhibiting PP2A activity and therefore promoting proliferation in HEK and leukaemia cells^7,18,45^. To determine whether SETBP1 interaction with SET is affected by variants, we performed co-immunoprecipitation assays in HEK293T/17 cells co-transfected with GFP-SET and FLAG-SETBP1 variants. We observed more abundant GFP-SET expression with increasing SETBP1 levels when co-expressed with FLAG-SETBP1 variants [p.(Leu957Pro) and p.(Thr962del)] compared to wild type (Figures 6C, 6D and S10B), consistent with the previously reported upregulation of SET with increasing SETBP1 levels^18^. We showed that mutated versions of FLAG-SETBP1, including those that led to faster fibroblast proliferation, retained interaction with GFP-SET similar to wild type (Figures 6C and 6D). Unexpectedly, we saw no differences from wild type for cell proliferation (Figure 6A) and interaction with SET (Figures 6C and 6D) in cells carrying a recurrent missense variant [p.(Glu858Lys)] which has also been reported in leukaemia cells in atypical chronic myeloid leukaemia (aCML) patients^7^. Moreover, we did not find any differences in *SET* expression between patient and control fibroblasts (Figure S10C), further suggesting that the aetiology involving identical variants in germline and somatic cells is likely to be cell-type specific. Overall, a subset of germline *SETBP1* variants outside the degron promote cell proliferation via a mechanism that is not driven by alterations in SET/SETBP1 interaction.

**Figure 6:**
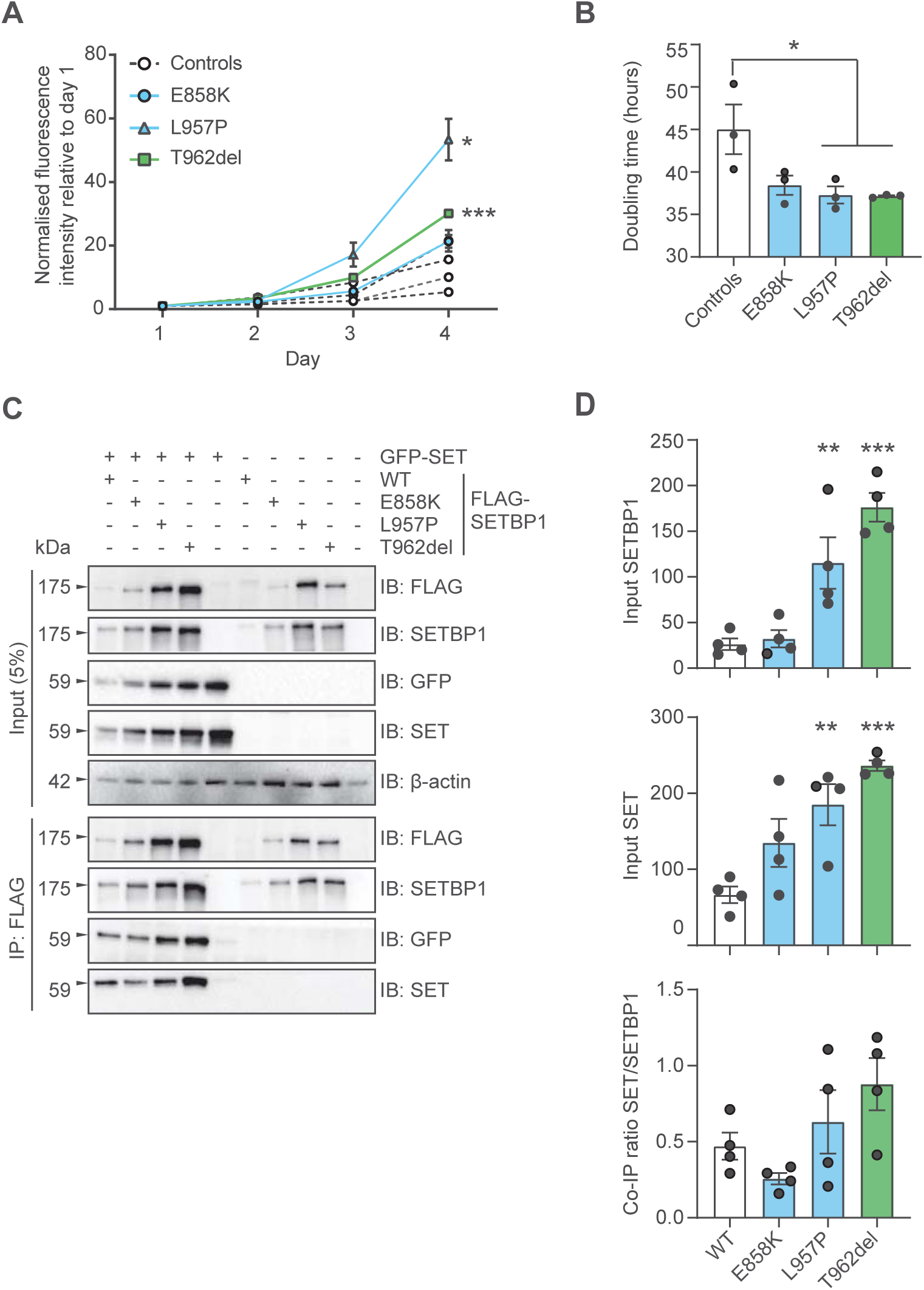
Increased proliferation of fibroblasts derived from patients carrying *SETBP1* variants outside the canonical degron. (A) CyQuant cell proliferation assay of fibroblasts from healthy individuals (controls) and patients carrying a *SETBP1* variant outside the degron. Nuclei of fibroblasts were stained with a GFP fluorescence dye. Fluorescence activity was measured daily for four days. Values are expressed relative to 1 day after seeding (day 1) and represent the mean ± SEM of three independent experiments, each performed in triplicate (\**p*<0.05, \*\*\**p*<0.001 versus controls; two-way ANOVA and a *post-hoc* Tukey’s test). (B) Cell doubling time of fibroblasts. Values represent the mean ± SEM of three independent experiments (\**p*<0.05 versus healthy controls; one-way ANOVA and a *post-hoc* Dunnett’s test). (C) Co-immunoprecipitation (Co-IP) was performed in whole cell lysates co-expressing FLAG-SETBP1 and GFP-SET. Wild type FLAG-SETBP1 and variants were co-immunoprecipitated using FLAG-conjugated magnetic agarose. Immunoblots were probed with an anti-FLAG, anti-SETBP1, anti-GFP or anti-SET antibody. β-actin was used as the loading control in the input fraction. (D) Quantification of SETBP1 (top) and SET (middle) levels in the input fraction. Quantification of SET co-immunoprecipitated with SETBP1 in the IP fraction was plotted (bottom). Values are expressed as the mean ± SEM of four independent experiments (\*\**p*<0.01, \*\*\**p*<0.001 versus WT; one-way ANOVA and a *post-hoc* Dunnett’s test).

### Patient fibroblasts carrying germline *de novo SETBP1* variants outside the degron show distinct transcriptomic profiles from healthy controls

To assess whether the observed *SETBP1* variants lead to a distinct gene expression signature compared to wild type, we performed RNA-seq on fibroblasts derived from three individuals carrying germline *SETBP1* variants outside the degron [p.(Glu858Lys) (female), p.(Leu957Pro) (male), and p.(Thr962del) (female)] as well as four healthy controls (two females and two males). Principal component analysis (PCA) revealed that transcriptomic profiles of patient fibroblasts did not cluster with those from healthy individuals (Figure 7A). We performed differential gene expression analysis comparing patient cells in females and males separately to controls. This analysis identified 4,110 differentially expressed genes (DEGs) in females (48.1% upregulated) and 2,479 DEGs in males (47.2% upregulated) (*p*<0.05, FDR) (Figure 7B, Tables S4 and S5). To further narrow down DEGs of interest, we used an intensity difference test, which assesses the statistical magnitude of effect, to find genes that showed unusually highly changing values^46^ in our samples. There were 218 (female) and 264 (male) DEGs that showed significant intensity differences (*p*<0.05, FDR) compared to healthy controls (Figure 7B, Tables S6 and S7). 32 DEGs (*p*<0.05, FDR) showed significant intensity differences (*p*<0.05, FDR) that were present in both female and male patient fibroblasts (Figures 7C and 7D, Table S8). Relative quantification of a subset of target genes (*BDKRB2*, *BRINP1*, *CHRM2*, *KRT19*, *RUNX3*, *SFRP2* and *LHX9*) by means of RT-qPCR confirmed the differential expression detected by RNA-seq (Figure 7E). We next performed gene ontology and gene set enrichment analyses (GSEA)^38,47^ of the consistent DEGs to delineate the most relevant biological pathways and cellular components. Functional annotation demonstrated over-representation (*p*<0.05, Benjamini-Hochberg FDR) of ontologies related to tissue development, cell proliferation and differentiation, cell surface receptor signalling, and response to stimuli (Figure 7F and Table S9). GSEA showed significant enrichment in patient samples of four gene sets including the G protein-coupled receptor signalling pathway, chemical homeostasis, ion transport and secretion (FDR < 25%; Figure S11E and Table S10). Cell proliferation was also found as one of the dysregulated pathways (Figures 7F and Table S9), consistent with the faster cell proliferation observed in patient fibroblasts (Figures 6A and 6B). Of note, enrichment of signalling pathways and cellular components comprising plasma membrane and vesicles was also detected in results of different GO analyses (Figures 7F and S11E). Six of the 32 differentially expressed genes (*ABCC9*, *ADGRG6*, *COMP*, *ELN*, *HLA-A,* and *MITF*) were present (significant overlap, *p*-value = 0.036, Fisher’s exact test) in the PanelApp database (intellectual disability v3.2 and autism v.0.22) (Figure S11A) showing their association with neurodevelopmental disorders including autism and intellectual disability. Nine of the genes (*RUNX3*, *SFRP2*, *CA12*, *SIM2*, *CPE*, *SCIN*, *FAM118A*, *ADGRG6,* and *PMEPA1*) were also dysregulated in HEK cells overexpressing an SGS variant [p.(Gly870Ser)] compared to empty controls, in a prior study^20^, suggesting a partial overlap with classical SGS pathology (Figure S11B). Furthermore, we cross-referenced these 32 DEGS with 70 putative SETBP1 target genes that were previously reported^20^ and identified *RUNX3,* a regulator of chromatin dynamics^48^, as a potential direct transcriptional target that was dysregulated in patient fibroblasts carrying *SETBP1* variants outside the degron (Figure S11C). Several direct transcriptional targets of SETBP1 (*MECOM, RUNX1, HOXA9, HOXA10* and *MYB*) have shown differential expression in leukaemia cells from aCML patients^20–23^ and in HEK cells overexpressing the p.(Gly870Ser) variant^20^. However, we did not observe differential expression of these genes in our RNA-seq data (Tables S4-8), further suggesting that the aetiological pathways are likely to be cell-type specific. Taken together, *SETBP1* variants outside the degron are associated with distinct transcriptomic profiles from healthy individuals and linked to ontologies related to tissue development, cellular signalling, transmembrane transport and membrane structure.

**Figure 7:**
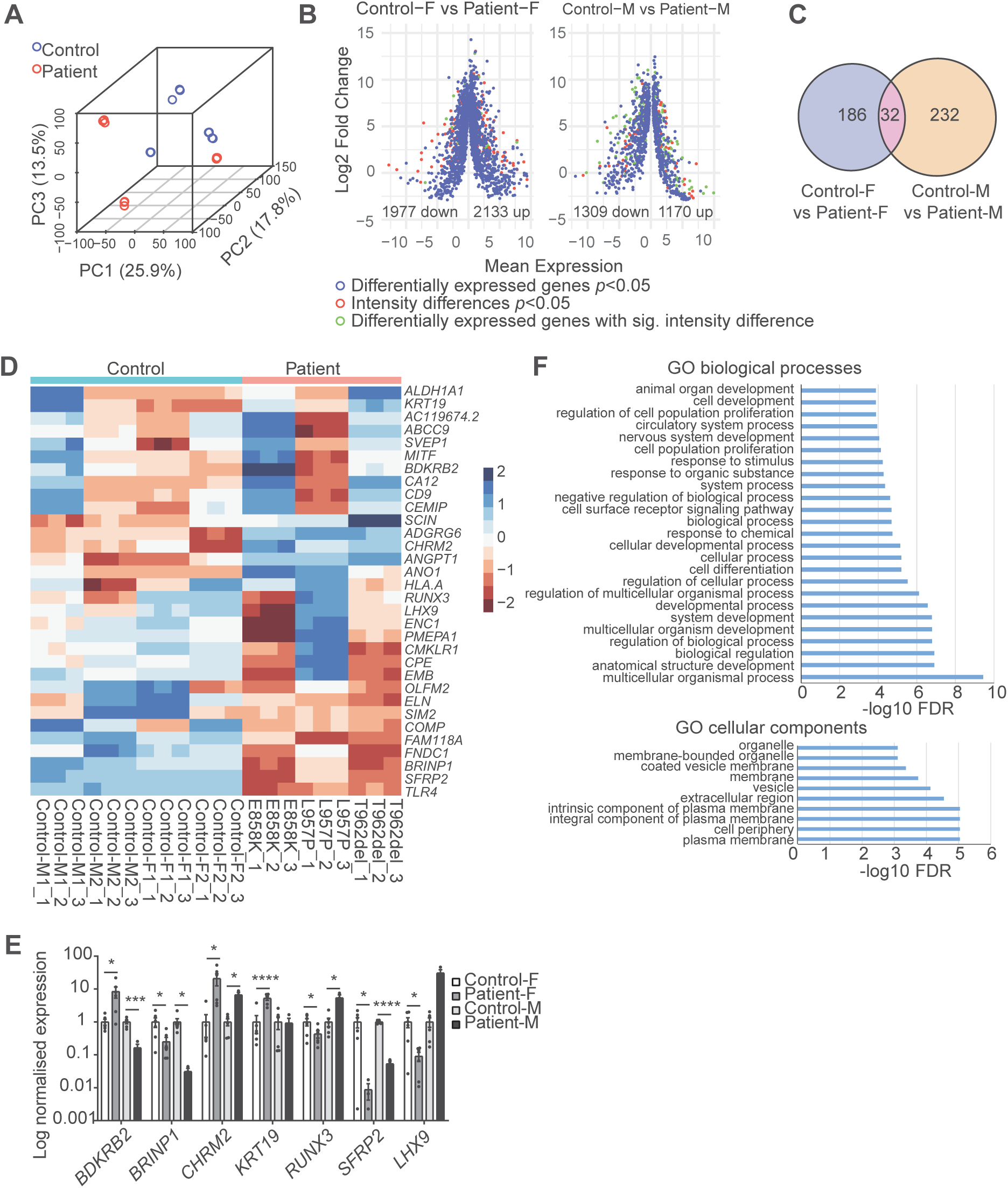
Fibroblasts carrying *SETBP1* variants showed distinct transcriptomic profiles from healthy controls. (A) Fibroblasts derived from patients harbouring a *SETBP1* variant (missense or in-frame deletion) outside the degron did not cluster with those from healthy individuals (controls). 3D principal component analysis (PCA) plot of variance distribution of four control fibroblast lines (blue; two females and two males), three patient lines (red; two females and one male); three technical replicates were included for each line. Principal component (PC) 1, PC2 and PC3 showed 25.9%, 17.8% and 13.5% of total variance respectively. (B) MA-plots showing the gene expression data of control vs. patient fibroblast lines (left: females; right: males) as a function of log ratios and mean average gene counts. (C) Venn diagram showing the overlap of genes that demonstrated significant differential expression (*p*<0.05, multiple testing correction with FDR) with significant intensity differences (adjusted *p*<0.05, multiple testing correction with FDR) in female control vs. female patient fibroblasts (left, blue) and male control vs. male patient fibroblasts (right, red). (D) Gene expression heatmap of the 32 DEGS with significant intensity differences in patient fibroblasts. (E) Validation of RNA-seq DEGs using RT-qPCR. Bars represent the mean ±SEM of three independent experiments (\**p*<0.05, \*\*\**p*<0.001, **** *p*<0.0001 versus female or male controls; one-way ANOVA and a *post-hoc* Dunnett’s test). (F, G) Dysregulated GO biological process and cellular components revealed by over-representation analysis of the 32 DEGs in patient fibroblasts using (F) over-representation analysis with g:Profiler (*p<*0.05, multiple testing correction with FDR).

## Discussion

Our study demonstrates that variant-specific functional follow-up is crucial to understand biological underpinnings of overlapping phenotypes and heterogeneity within cohorts. Work from our group and others have previously delineated the phenotypic heterogeneity within cohorts carrying germline *SETBP1* loss-of-function variants causing *SETBP1* haploinsufficiency disorder, which have much milder clinical correlates than classical SGS^6,15,16^. Here, we present, to our knowledge, the largest cohort of individuals who have *SETBP1* variants (missense and in-frame deletion) outside the canonical degron region, with clinical features that do not fit with the original diagnostic criteria of classical SGS. We used a genotype-driven approach combining deep clinical and speech phenotyping with functional follow-up of specific variants to investigate the phenotype-genotype associations and delineate the underlying pathophysiological mechanisms. Our results show that individuals carrying variants outside the degron display a broad spectrum of clinical features of variable severity that only partially overlap with either SGS or *SETBP1* haploinsufficiency disorder. This wide clinical spectrum could be explained by the heterogeneous pathophysiological mechanisms resulting from these genetic variations, as shown in the cell-based experiments. Using an array of functional assays, we showed that variants outside the degron disrupt SETBP1 protein functions via aetiological mechanisms including impairments in ubiquitination, DNA binding capacity, transcription and cell proliferation, which are independent of SETBP1 protein levels. Interestingly, we found that *SETBP1* variants led to reduced transcription of its direct target *FOXP2,* of which rare variants are a known cause of monogenic speech disorder characterised by CAS^42–44^.

### Variable severity of broad clinical features depends on proximity of variants to the degron

The variable severity of the broad spectrum of clinical features observed in our cohort could largely be categorised into three groups based on the distance of the variant from the canonical degron. We found that patients with variants in close proximity to the degron within the SKI domain (affecting amino acids 862-874, excluding the degron 868-871) showed the most severe phenotype with severe intellectual disability, inability to speak and are often unable to walk. These individuals are much more severely affected than those with *SETBP1* haploinsufficiency disorder^16^ and yet do not fulfil the original diagnostic criteria of classical SGS as proposed by Lehman *et al*.^41^, which were proposed at a time when the causative gene was not identified and may have led to an ascertainment bias in the first studies after identification of the gene^2,6^. In 2020, Sullivan *et al.*^49^ described an individual with a SETBP1 variant affecting amino acid 871 within the hotspot region, who displayed an atypical milder phenotype which was not concordant with the original diagnostic criteria of SGS. She had a moderate intellectual disability, no congenital anomalies, and showed less apparent dysmorphisms than patients with classical SGS including mild midface retraction, hypertelorism, short upturned nose and prominent low-set ears. This more mildly affected individual than those with classical SGS may point to a broader SGS-associated phenotype than originally defined, with a different (non-lethal) life expectancy, possibly involving interactions with other variants in the genomic background. In addition, this wider spectrum of SGS may also be applicable to cases with aetiological mutations in the 862-874 region who displayed similar facial features including mild midface retraction, microcephaly and prominent low-set ears.

The group of *SETBP1* variants affecting residues at positions 854-858 (located further from the degron) showed a phenotype including mild to moderate intellectual disability. Motor development was delayed but they all achieved the milestone to walk without support. Growth was normal and we did not observe a consistent pattern of dysmorphisms, but this may be partly because we had only a limited number of patient photographs, and these came from different age ranges. Future studies including a larger cohort whose facial features are examined at standardised age ranges longitudinally (with photographs taken from different angles) will be beneficial to profiling any dysmorphism and thus improving facial feature-based clinical diagnosis. The p.(Glu858Lys) variant was noted in four individuals. Leonardi *et al.*^50^ also described an individual with the same variant, showing severe intellectual disability and epilepsy. In that study, the variant was reported to be inherited from an asymptomatic mother whose variant was *de novo*. Unfortunately, no other tissues were tested in the mother to exclude the possibility of mosaicism.

Individuals with variants located outside the SKI domain (n=4) showed a more variable clinical phenotype ranging from mild to severe intellectual disability. Two of these individuals [carrying p.(Leu957Pro) and p.(Thr962del) variants] showed a very similar facial phenotype including a round face, blepharophimosis, hypertelorism and a short nose with a bulbous tip, features also often noted in individuals with *SETBP1* haploinsufficiency disorder. The two individuals with variants located furthest from the SKI domain [p.(Ser444Arg) and p.(Val657Ala)] had inherited them from affected mothers who also have low IQ (grandparents and other family members were not available for testing). We therefore also included these variants in the functional cellular assays together with the *de novo* variants outside the degron in our study. Indeed, these two variants showed significant functional impacts on the SETBP1 protein as shown in our cellular assays. In light of this, we recommend that clinicians should examine the clinical phenotypes of family members carrying the same variants as well as performing cellular assays to test pathogenicity when assessing pathogenicity of inherited variants of uncertain significance based on ACMG (American College of Medical Genetics and Genomics) guidelines^51^. Clinicians could also consider performing functional prediction using algorithms where available, such as those for known epilepsy ion channel genes^52^.

The variable phenotypic severity and partial clinical overlaps with SGS or *SETBP1* haploinsufficiency disorder suggest the potential existence of a third clinical entity involving disruptions of this gene. Alternatively, different types of *SETBP1* variants could lead to a continuum of clinical features of SETBP1-related disorders with SGS and *SETBP1* haploinsufficiency disorder positioned at two extremes of severity. Investigations of future larger cohorts including individuals with different variants in combination with genome-wide analysis, for example, DNA methylation analysis (episignatures)^53,54^, will provide clarification over the clinical definition and aetiology, as well as improving clinical diagnosis. Our participants had long-term chronic difficulties across a number of developmental domains. Individuals with *SETBP1* variants outside the degron region should receive careful assessment across core domains of speech and language, attention, motor and sleep as early as possible following diagnosis, leading to better targeted and earlier intervention to optimise children’s health and developmental outcomes.

### *FOXP2* is a direct transcriptional target of SETBP1

Individuals with *SETBP1* haploinsufficiency disorder showed significant impairments in both receptive and expressive language, suggesting SETBP1 as a strong candidate for speech and language disorders^15^. Of note, we have here demonstrated that individuals carrying *SETBP1* variants outside the degron, where speech and language data were available, showed generally low language ability for all subdomains including expressive, receptive, written and social language. Intriguingly, our cell-based experiments identified *FOXP2* as a novel direct transcriptional target of SETBP1 (Figure 5C). All functionally tested *SETBP1* variants outside the degron displayed significantly decreased *FOXP2* transcription in luciferase reporter assays. *FOXP2* is one of the few genes identified to-date for which disruptions yield disproportionate effects on speech and language, yet there is little known about its upstream regulators. Our results provide molecular evidence linking two strong candidate genes implicated in childhood apraxia of speech, which might help provide insight into the severe speech and language impairments observed in our, and other, cohorts. We did not detect *FOXP2* as a differentially expressed gene in our transcriptomic analysis of patient fibroblasts or in those performed on patient leukaemia cells and cell lines expressing *SETBP1* constructs in previous studies, further pointing towards potential cell type-specific aetiological pathways.

### A broad range of functions are disrupted by variants outside the degron via pathophysiological mechanisms independent of SETBP1 protein dosage

As seen in the summary of our functional analyses (Figure 8), a broad spectrum of cellular functions was affected. In these cellular assays, variants outside the degron led to molecular consequences distinct from classical SGS variants. Classical SGS variants showed increased protein stability and higher SETBP1 protein levels as a result of impaired degradation by proteasome and autophagy. Moreover, while the binding capacity of SGS variants to AT-rich DNA sequences was not affected, a subset of SGS variants showed higher transcriptional activation, suggesting gain of function. These findings are consistent with prior studies showing global upregulation of SETBP1 binding to genomic regions and increased chromatin accessibility when overexpressing an SGS variant [p.(Gly870Ser)] in HEK cells^20^.

**Figure 8:**
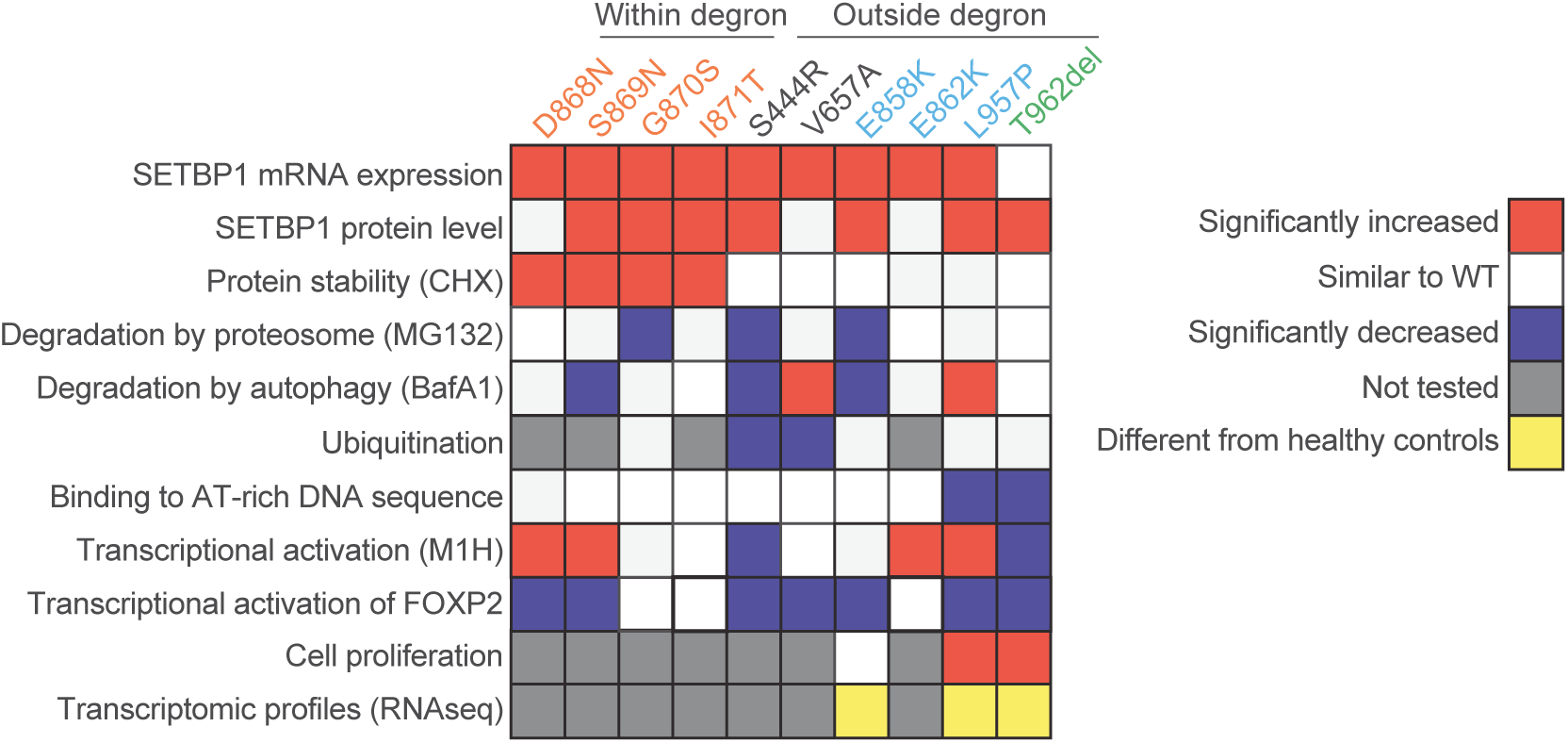
Summary of functional assays: variants outside the degron disrupt SETBP1 functions by dysregulating transcription, reducing DNA binding capacity and promoting cell proliferation independent of SETBP1 level. A heat map summarising the functional characterisation of *SETBP1* variants within (classical SGS) and outside the degron. Classical SGS variants showed increased protein stability and higher SETBP1 protein levels. While binding capacity of SGS variants to AT-rich DNA sequences was not affected, a subset showed higher transcriptional activation, suggesting a gain of function. In contrast, *SETBP1* variants outside the degron demonstrated a broad spectrum of functional disruptions which could be largely categorised into two groups. Although the majority of these variants resulted in more abundant SETBP1 protein, there were variable degrees of disruption in SETBP1 degradation via the proteasome and autophagy machinery. Variants furthest from the degron (grey) showed mostly reduced SETBP1 degradation by both proteasome and autophagy due to disrupted ubiquitination, while only a subset of those in the proximity of the degron demonstrated impaired degradation and mildly reduced ubiquitination. Two variants outside the degron led to lower affinity to AT-rich DNA sequences, suggesting loss of function. Transcriptional activation was affected in the majority of variants outside the degron to various degrees. Increased proliferation was seen for patient fibroblasts carrying two variants outside the degron, suggesting a partially overlapping mechanism with classical SGS. Patient fibroblasts carrying variants outside the degron showed distinct transcriptomic profiles from healthy controls, implicating biological pathways involved in system/tissue development, cell proliferation and differentiation, cell surface receptor signalling, and membrane composition

In contrast, *SETBP1* variants outside the degron demonstrated an array of functional disruptions that only partially overlapped with those observed for SGS variants. Although the majority of variants outside the degron resulted in more abundant SETBP1, there were variable extents of disruption in degradation of this protein via the proteasome and autophagy machinery, suggesting that impaired protein degradation might not be a core pathogenic mechanism. The p.(Glu862Lys) variant that is in close proximity to the degron led to increased protein levels and transcriptional activation, similar to SGS variants, but neither protein stability nor degradation were affected. This points to a functional landscape partially overlapping with SGS variants but distinct from the other variants located further from the degron. The variant further away from the degron but still in the SKI domain [p.(Glu858Lys)] led to more abundant SETBP1 protein, reduced degradation via proteasome and autophagy and lower *FOXP2* transcription. However, even though p.(Glu858Lys) showed functional results overlapping with those of SGS variants, the individual carrying the variant still presented milder clinical features compared to classical SGS (amino acids 868-871) and to those with variants nearest to the degron (amino acids 862-867 and 873-874).

Interestingly, functional impairments of variants further away from SKI domain could be largely divided into two groups, matching with the variable clinical features observed in the individuals that carried them. In our cell-based assays, two variants [p.(Leu957Pro) and p.(Thr962del)] led to a loss of DNA-binding and transcriptional activity despite the increased protein levels detected in transfected cells and patient fibroblasts, consistent with the observed clinical features being closer to individuals with *SETBP1* haploinsufficiency disorder. These two variants are located close to the HCF binding site (amino acids 991–994), which is an important component of the COMPASS complex. Piazza *et al.* reported that mutant SETBP1 still retained the ability to bind to HCF, PHF6/8 and KMT2A in HEK cells overexpressing an SGS variant [p.(Gly870Ser)]^20^. This raises a hypothesis that the loss of transcriptional activity for variants such as p.(Leu957Pro) and p.(Thr962del) could result from disruption of interaction with HCF, a core protein of the SET1/KMT2A COMPASS-like complex responsible for H3K4 mono- and di-methylation, and chromatin accessibility^20,55^. In particular, the threonine residue at the 962 position is likely crucial for SETBP1 transcriptional activity, potentially via HCF1-related pathways given its proximity to the HCF-binding site and the complete loss of transcriptional activation in cells expressing p.(Thr962del). Intriguingly, variants that were furthest from the degron region [p.(Ser444Arg) and p.(Val657Ala)] showed mostly impaired SETBP1 degradation by both proteasome and autophagy due to disrupted ubiquitination. It is possible that these variants are physically closer to the degron in the three-dimensional conformation of the protein. However, we could not predict how *SETBP1* variants might affect its protein folding or interactions with interactors due to limited knowledge about the overall structure of the protein.

SETBP1 has been identified as a protein with intrinsically disordered regions (IDRs), which are regions that are prone to mutations and are often found in proteins associated with cancers^56^. Indeed, the majority of *SETBP1* missense variants identified to-date, both somatic and germline, are located within an IDR (amino acids 858-880)^56^. Many proteins containing IDRs do not have a single well-defined conformation, and the conformation changes depending on interactions with cofactors. Most proteins with IDRs that are elements of the transcriptional machinery are multifunctional, involved in processes such as regulating degradation via a linear motif, binding to genomic DNA, activating/repressing transcription, and/or modulating histone modifications^56^. This wide range of functions could explain the broad spectrum of impairments found in our cell-based assays and make it more difficult to predict how variants affect SETBP1 function. Future studies that profile and delineate the structural impact of *SETBP1* variants and how they affect interactions with cofactors will aid the understanding of their impacts on protein functions and thus aetiology.

### Cell-type specific aetiology of *SETBP1* variants

We identified a subset of differentially expressed genes in our RNA-seq data that also showed differential expression in prior analyses of p.(Gly870Ser) expressing cells^20^, suggesting partly shared aetiological pathways with SGS, complementing the observed partial overlap in clinical manifestation. Nevertheless, we did not observe overlaps with genes dysregulated in aCML patient cells, nor did we observe any changes in proliferation in fibroblasts derived from individuals carrying recurrent variant p.(Glu858Lys) that has also been reported in individuals with leukaemia^7^. This is consistent with the findings of a recent study where disruption of SET/PP2A pathway was not detected in SGS neural progenitors and cortical organoids^8^, suggesting that the aetiological pathways underlying germline and somatic *SETBP1* variants are likely cell type-/tissue-specific. Future studies investigating the spatial and temporal expression of SETBP1 and the functional impact of variants during brain development are much needed to understand the pathways that go awry. This could potentially be achieved by using brain organoids that carry *SETBP1* variants. A recent study investigating SGS variants in young cortical organoids and neural progenitors suggested p53-related pathways as a novel mechanism involved in this disorder^8^. *SETBP1* knockout neural progenitors showed protracted proliferation and distorted layer-specific neuronal differentiation with overall decrease in neurogenesis via WNT/β-catenin signaling pathway in a recent BioRxiv preprint^24^. These investigations focused on relatively young neural progenitors as well as either an SGS variant or gene deletions. Future work comparing the functional impacts of different types of variants in neuronal models is essential to delineate the aetiology, complexity and pleiotropy of SETBP1-related disorders.

### SETBP1 is a central upstream element of biological networks disturbed in neurodevelopmental disorders

Our RNA-seq analysis of patient fibroblasts carrying variants outside the degron revealed a number of DEGs including transcription factors that are important for neurodevelopment and associated with intellectual disability and/or autism (Figure S11D). Previous chromatin immunoprecipitation and mass spectrometry analyses in HEK cells overexpressing SETBP1 have shown its role as a potential epigenetic hub^20^. Moreover, SETBP1 directly regulates transcription of *FOXP2*, which has been shown to interact with an array of transcription factors implicated in neurodevelopmental disorders characterised by speech and language deficits^57,58^. Given the broad expression of SETBP1 in neural progenitors and neurons, and its roles as a chromatin remodeller and transcription factor, our findings further suggest that this protein is a central element of biological networks regulating brain development, and could be important in pathophysiology of neurodevelopmental disorders caused by genetic disruptions of its interactors or downstream targets.

In conclusion, in this study, we investigated the genotype-phenotype associations of germline *SETBP1* variants by thoroughly profiling the clinical and speech-language phenotypes. We have established an array of functional assays for testing pathogenicity of the identified variants, which can be applied to variants of uncertain significance in the future. Despite the partial overlap of results in clinical and functional analyses with SGS, our data suggest that *SETBP1* variants outside the degron cause a clinically and functionally variable form of neurodevelopmental disorder that is milder than SGS and in certain aspects more severe than *SETBP1* haploinsufficiency disorder. In particular, we have shown impairments in ubiquitination, DNA binding capacity, transcription and cell proliferation depending on the proximity of variants relative to the degron in the SKI domain. Overall, our findings reveal that variants outside the degron act via a range of loss-of-function pathophysiological mechanisms independent of protein abundance, unlike SGS and *SETBP1* haploinsufficiency disorder, providing valuable new insights into diagnosis and aetiology of SETBP1-related disorders.

## Supplemental Data

Supplemental Data include 11 figures and 12 tables.

## Declaration of Interests

The authors declare no other competing interests.

## Data Availability

All data produced in the present study are available upon reasonable request to the authors

## Acknowledgments

We are grateful to all individuals and families for their contribution. We would like to thank the members of the Cell Culture Facility, Department of Human Genetics, Radboud university medical center, Nijmegen, for cell culture of proband-derived cell lines. We especially thank Joery den Hoed, Else Eising (Language and Genetics Department, MPI for Psycholinguistics, Nijmegen), Alexander Hoischen, Lot Snijders-Blok (Department of Human Genetics, Radboud university medical center, Nijmegen) and Rocio Acuna-Hidalgo (Nostos Genomics, Berlin) for scientific discussion. This work was supported by the Max Planck Society (M.M.K.W., R.A.K., G.A., A.V., and S.E.F.). W.K.C was funded by SFARI and the JPB Foundation. M.S.H., I.E.S., and A.T.M. were funded by a National Health and Medical Research Council (NHMRC) Centre of Research Excellence Grant (1116976), an Australian Research Council (ARC) Discovery Project (DP120100285), an NHMRC Project Grant (1127144), and the March of Dimes Grant Scheme. M.S.H. was funded by an NHMRC Career Development Fellowship (1063799). I.E.S. was funded by an NHMRC Investigator Grant (1172897), an NHMRC Practitioner Fellowship (1006110), and NHMRC Development Grant (1153614). A.T.M. was funded by an NHMRC Investigator Grant (1195955), an NHMRC Practitioner Fellowship (1105008), and an NHMRC Development Grant (1153614). E.P. was supported by the National Health and Medical Research Council (GNT11149630), Australia and the research WGS was supported by NHMRC.

## Web Resources

CADD, https://cadd.gs.washington.edu

gnomAD, https://gnomad.broadinstitute.org

MetaDome, https://stuart.radboudumc.nl/metadome

OMIM, https://www.omim.org

UniProt, https://www.uniprot.org

Spatial Clustering, https://github.com/laurensvdwiel/SpatialClustering

g:GOSt (part of g:Profiler), https://biit.cs.ut.ee/gprofiler/gost

The Gene Ontology Resource, http://geneontology.org/docs/go-enrichment-analysis/

COSMIC: https://cancer.sanger.ac.uk/cosmic

Seqmonk: https://www.bioinformatics.babraham.ac.uk/projects/seqmonk/

## Data and Code Availability

Code used in the spatial clustering analysis is available at: https://github.com/laurensvdwiel/SpatialClustering^33^. Codes of RNA-seq data analysis are available on request.

## Permissions

No permission was needed.

## Notes

### Competing Interest Statement

The authors have declared no competing interest.

### Author Declarations

Ethics committee/IRB of Commissie Mensgebonden Onderzoek Regio Arnhem-Nijmegen under number 2011/188 gave ethical approval for this work The Human Research Ethics Committee of The Royal Children's Hospital, Melbourne, Australia, approved study and testing on a research basis of proband 1 (Project 37353).

