## Supplemental information and Supplementary Tables S11-S12 for "*SETBP1* variants outside the degron disrupt DNA-binding and transcription independent of protein abundance to cause a heterogeneous neurodevelopmental disorder"

**Figure S1: Detailed clinical features.** Not available in this preprint

**Figure S2: MetaDome analysis of the *SETBP1* missense variants.**

**Figure S3: Frequency of somatic and germline *SETBP1* variants.**

**Figure S4: SETBP1 expression in transiently transfected HEK293T/17 cells.**

**Figure S5: Protein stability and subcellular localization of SETBP1 protein in human dermal fibroblasts.**

**Figure S6: Relative expression SETBP1 variants as YFP-fusion protein in HEK293T/17 cells treated with 50µg/mL cycloheximide (CHX) or DMSO vehicle control.**

**Figure S7: Relative expression SETBP1 variants as YFP-fusion protein in HEK293T/17 cells treated with 5µg/mL MG132 or DMSO vehicle control.**

**Figure S8: Relative expression SETBP1 variants as YFP-fusion protein in HEK293T/17 cells treated with 100nM Bafilomycin (BafA1) or DMSO vehicle control.**

**Figure S9: SETBP1 affinity to two AT-rich DNA consensus sequences.**

**Figure S10: Proliferation of patient fibroblasts carrying *SETBP1* variants outside the degron and SETBP1 interaction with SET.**

**Figure S11: Venn diagram of differentially expressed genes in fibroblasts.**

**Table S1: Clinical information per individual.** Not available in this preprint

**Table S2: List of variants, including annotations and classification based on ACMG guidelines.**

**Table S3: Frequency of variants in cancer.**

**Table S4: differentially expressed genes in female patient HDF compared to female control HDF (DESeq2 hits).**

**Table S5: differentially expressed genes in male patient HDF compared to male control HDF (DESeq2 hits).**

**Table S6: differentially expressed genes in female patient HDF compared to female control HDF (DESeq2 hits) with intensity differences (*p*<0.05 after multiple testing correction).**

**Table S7: differentially expressed genes in male patient HDF compared to male control HDF (DESeq2 hits) with intensity differences (*p*<0.05 after multiple testing correction).**

**Table S8: differentially expressed genes with intensity differences (*p*<0.05 after multiple testing correction) in both female and male patient HDF (32 genes).**

**Table S9: List of genes in significant GO-terms using g:Profiler.**

**Table S10: List of enriched gene sets in GSEA.**

**Table S11: List of primers.**

| **Name** | **Sequence (5’-3’)** | **Application** |
| --- | --- | --- |
| SETBP1 S444R F | CCTGTTAACTACTTCACTCCTCATGGTGATTCCAGAA | SDM |
| SETBP1 S444R R | TTCTGGAATCACCATGAGGAGTGAAGTAGTTAACAGG | SDM |
| SETBP1 V657A F | GGTCTTCTTATCCAACGCGCCGAGCTTTCCAA | SDM |
| SETBP1 V657A R | GTTGGAAAGCTCGGCGCGTTGGATAAGAAGACC | SDM |
| SETBP1 E858K F | CTCACTGTGGGACTTGCTCACAGGGGACA | SDM |
| SETBP1 E858K R | TGTCCCCTGTGAGCAAGTCCCACAGTGAG | SDM |
| SETBP1 L957P F | GGTGATTAGCTCCTCCGGGTCTGCCAGAAACTG | SDM |
| SETBP1 L957P R | CAGTTTCTGGCAGACCCGGAGGAGCTAATCACC | SDM |
| SETBP1 T962del F | AACACTTGGAACTTGATTAGCTCCTCCAGGTCTGC | SDM |
| SETBP1 T962del R | GCAGACCTGGAGGAGCTAATCAAGTTCCAAGTGTT | SDM |
| 5’-AAAATAA-3’ S | Twelve repeats of 5’-AAAATAA-3’ | Cloning |
| 5’-AAAATAA-3’ AS | Twelve repeats of 5’-TTTTATT-3’ | Cloning |
| 5’-AAAATAT-3’ S | Twelve repeats of 5’-AAAATAT-3’ | Cloning |
| 5’-TTTTATA-3’ AS | Twelve repeats of 5’-TTTTATA-3’ | Cloning |
| ALDH1A1 F | TCGTCTGCTGCTGGCGACAA | RT-qPCR |
| ALDH1A1 R | AGCCCAACCTGCACAGTAGCG | RT-qPCR |
| ANO1 F | GAAAATCCATGCCCCCTGGA | RT-qPCR |
| ANO1 R | CACTTTGGGCTGGATGGGAT | RT-qPCR |
| BDKRB2 F | CTGTCTGTTCGTGAGGACTCC | RT-qPCR |
| BDKRB2 R | CTGGATGGTGTTGAGCCAGC | RT-qPCR |
| BRINP1 F | ATCATGGAGTACACGCTGGC | RT-qPCR |
| BRINP1 R | ATTGCCCCAGTGCTGATGG | RT-qPCR |
| CHRM2 F | ATGCTGCTGTCACCTTTGGT | RT-qPCR |
| CHRM2 R | AGAAACGGGGTCTTGGTTGG | RT-qPCR |
| ELN F | GCAGGAGTTAAGCCCAAGG | RT-qPCR |
| ELN R | TGTAGGGCAGTCCATAGCCA | RT-qPCR |
| EMB F | CTTGTTTCTTTCGAGAGGAAAAGGAAC | RT-qPCR |
| EMB R | TTACATGTCAAGACAGTAGAATCCCC | RT-qPCR |
| HLA-A F | AAAAGGAGGGAGTTACACTCAGG | RT-qPCR |
| HLA-A R | GCTGTGAGGGACACATCAGAG | RT-qPCR |
| HOXA9 F | CCACGCTTGACACTCACACT | RT-qPCR |
| HOXA9 R | AGTTGGCTGCTGGGTTATTG | RT-qPCR |
| HOXA10 F | CTCGCCGGAGAAGGATTC | RT-qPCR |
| HOXA10 R | AGTTTCATCCTGCGGTTCTG | RT-qPCR |
| HPRT1 F | AGATGGTCAAGGTCGCAAG | RT-qPCR |
| HPRT1 R | GTATTCATTATAGTCAAGGGCATATCC | RT-qPCR |
| KRT19 F | CCACTACTACACGACCATCCA | RT-qPCR |
| KRT19 R | AGGACAATCCTGGAGTTCTCAA | RT-qPCR |
| LHX9 F | CTTTGCCAAGGACGGTAGCA | RT-qPCR |
| LHX9 R | CAGGTGAAGCAGCTCAGGTG | RT-qPCR |
| MECOM F | GAGAGCAGCCTTACAGATGCA | RT-qPCR |
| MECOM R | CGATGTTGCTGTACCGGACA | RT-qPCR |
| MYB F | GAAAGCGTCACTTGGGGAAAA | RT-qPCR |
| MYB R | TGTTCGATTCGGGAGATAATTGG | RT-qPCR |
| RUNX1 F | CCCTAGGGGATGTTCCAGAT | RT-qPCR |
| RUNX1 R | TGATGGCTCTGTGGTAGGTG | RT-qPCR |
| RUNX3 F | GACAGCCCCAACTTCCTCT | RT-qPCR |
| RUNX3 R | CACAGTCACCACCGTACCAT | RT-qPCR |
| SCIN F | TCTGCGTTCCTGACTGTTC | RT-qPCR |
| SCIN R | GACCTCCTTTCTTTGATGTTCC | RT-qPCR |
| SET F | TCTGGAAAGGATTTGACGAAACG | RT-qPCR |
| SET R | GGTTCCTCATGCTGCCTCTT | RT-qPCR |
| SETBP1 F | CTCCTCCATGTCTCCAGGGAT | RT-qPCR |
| SETBP1 R | GGCACTGAGCCTTCGTTCTT | RT-qPCR |
| SFRP2 F | ATGACAACGACATAATGGAAACGC | RT-qPCR |
| SFRP2 R | GCTCTTGGTCTCCAGGATGATTT | RT-qPCR |
| SIM2 F | AGACCCTATACCATCACGTGCA | RT-qPCR |
| SIM2 R | CTTGGACAGCAGCCGGTAGTA | RT-qPCR |
| TBP F | GGGCACCACTCCACTGTATC | RT-qPCR |
| TBP R | CGAAGTGCAATGGTCTTTAGG | RT-qPCR |

Sequences of primers and oligos used in the current study. F=forward primer; R=reverse primer; S=sense; AS=anti-sense; SDM=Site-Directed Mutagenesis.

**Table S12: List of antibodies.**

| **Target** | **Host** | **Company** | **Cat. no** | **Dilution** | **Application** |
| --- | --- | --- | --- | --- | --- |
| Primary antibodies | | | | | |
| β-actin | Ms | Sigma-Aldrich | A5441 | 1:10000 | IB |
| DKK/FLAG | Ms | Origene | TA50011-100 | 1:2000 | IB |
| DKK/FLAG | Ms | Origene | TA50011-100 | 1:1000 | IF |
| JL-8 | Ms | Clonetech | 632380 | 1:8000 | IB |
| Ki-67 | Ck | EnCor Biotech | CPCA-Ki67 | 1:500 | IF |
| MCM2 | Rb | Abcam | ab108935 | 1:20000 | IB |
| PCNA | Ms | SantaCruz | Sc-56 | 1:200 | IB |
| SET | Rb | Abcam | ab1183 | 1:10000 | IB |
| SETBP1 | Rb | ProteinTech | 16841-1-AP | 1:10000 | IB |
| SETBP1 | Rb | Atlas Antibodies | HPA049022 | 1:200 | IF |
| SUMO2/3 | Rb | Abcam | Ab9132 | 1:1000 | IB |
| Ubiquitin | Rb | Cell Signalling | 43124S | 1:1000 | IB |
| Seconady Antibodies | | | | | |
| AlexaFluor 488-conjugated-anti-rabbit IgG | Gt | Invitrogen | A11034 | 1:1000 | IF |
| AlexaFluor 568-conjugated-anti-mouse IgG | Gt | Invitrogen | A11031 | 1:1000 | IF |
| AlexaFluor 568-conjugated-anti-rabbit IgG | Gt | Invitrogen | A11036 | 1:1000 | IF |
| AlexaFluor 647-conjugated-anti-chicken IgG | Gt | Invitrogen | A21449 | 1:1000 | IF |
| HRP-conjugated-anti-rabbit IgG | Gt | Jackson ImmunoResearch | 111-035-003 | 1:10000 | IB |
| HRP-conjugated-anti-mouse IgG | Gt | Jackson ImmunoResearch | 115-007-003 | 1:10000 | IB |

Ms= mouse; Ck=chicken; Rb=rabbit; Gt=goat; IB=immunoblotting; IF=immunofluorescence
