## Supplementary Figures S1-S11 for "*SETBP1* variants outside the degron disrupt DNA-binding and transcription independent of protein abundance to cause a heterogeneous neurodevelopmental disorder"

#### Figure S2: MetaDome analysis of the SETBP1 missense variants.

(A) Overview of the SETBP1 protein (transcript NM_015559.2, ENST00000282030.5) intolerance landscape visualized via the MetaDome web server version 1.0.1. The tolerance landscape is computed based on single nucleotide variants present in the gnomAD database. It is calculated as a missense over synonymous ratio in a sliding window of 21 residues over the entire SETBP1 protein. The green and blue peaks correspond to regions more tolerant to missense variation, and the red valleys indicate intolerant regions. The locations of the variants in our cohort are displayed within the tolerance landscape of SETBP1. Most of these variants are predicted to be intolerant to highly intolerant to missense variation. Previously reported SGS variants located within the degron were also indicated. (B) Detailed overview of the intolerance landscape of the SETBP1 SKI domain where missense variants cluster. Canonical degron is highlighted.

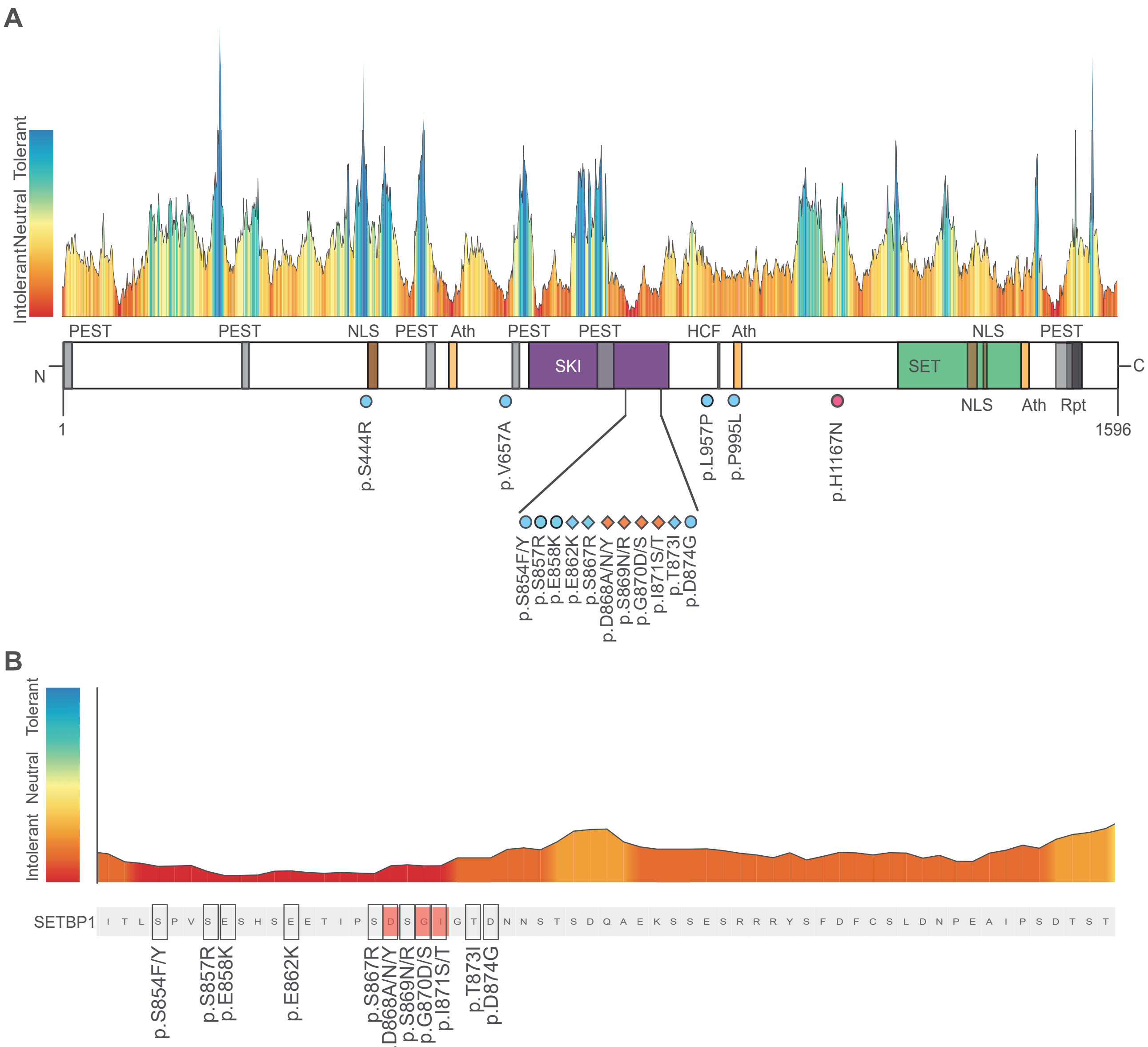

### Figure S3: Frequency of somatic and germline SETBP1 variants.

Frequency of all SETBP1 missense variants in (A) all tissues and (B) haematopoietic and lymphoid cells. (C) Frequency and (D) proportion of germline vs somatic missense variants included in this study.

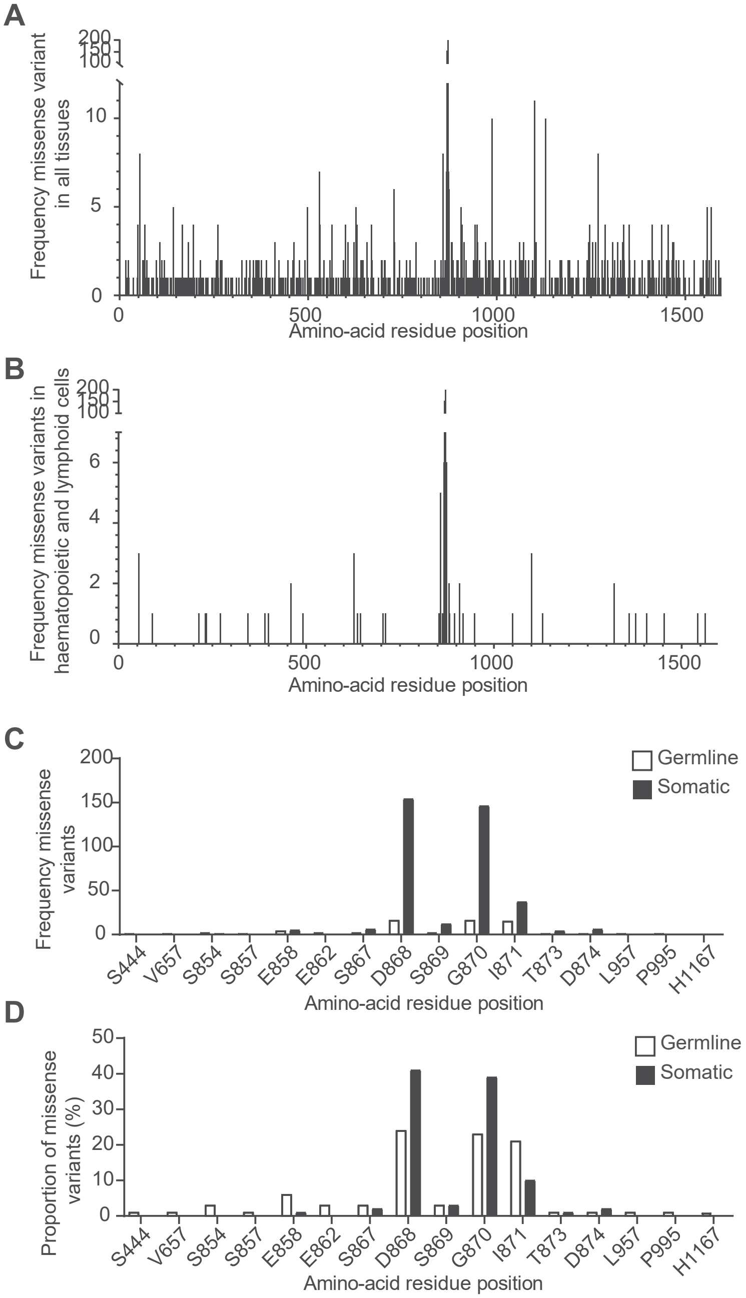

#### Figure S4: SETBP1 expression in transiently transfected HEK293T/17 cells.

(A) Immunoblot of whole cell lysates of HEK293T/17 cells expressing YFP-tagged SETBP1 variants probed with anti-SETBP1 and anti-GFP antibodies. β-actin was used as a loading control. Representative blots of three independent experiments are shown. (B) Immunoblot of whole cell lysates of HEK293T/17 cells expressing GAL4-fused SETBP1 variants probed with anti-SETBP1 antibody. (C) Confocal microscopy images of SETBP1 (green) and FLAG (red) localization of WT FLAG-SETBP1 and variants. Results are representative of three independent experiments. Scale bar= 5µm. (D) Direct fluorescence imaging of cells expressing YFP-tagged variants of the SETBP1 protein using confocal microscopy. Wildtype and all variants show a speckle-like pattern in the nucleus. Nuclei are stained with Hoechst 33342 (blue). Scale bars= 5µm .

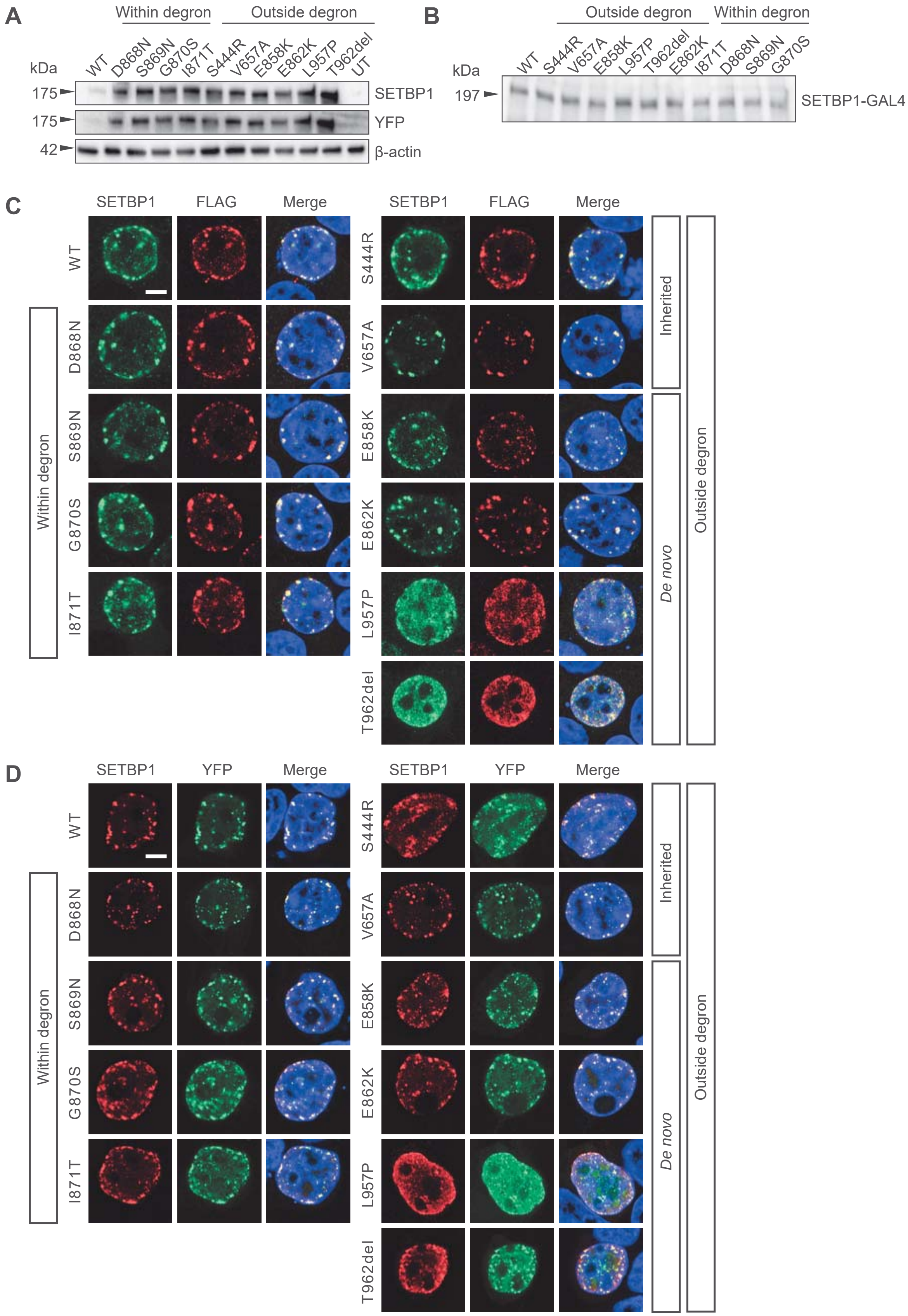

#### Figure S5: Protein stability and subcellular localization of SETBP1 protein in human dermal fibroblasts.

(A) Fibroblasts derived from healthy controls and patients carrying SETBP1 variants outside the degron were treated with a translation inhibitor cycloheximide (CHX; 100µg/ml) or vehicle control DMSO for 4 hours. Immunoblots of whole cell lysates probed with an anti-SETBP1 antibody were shown. B-actin was used a loading control. Results are representative of three independent experiments. (B) Confocal microscopy images of immunostaining of SETBP1 (green). Nuclei were stained with Hoechst 33342 (blue). Wildtype and all variants show a speckle-like pattern in the nucleus. Results are representative of three independent experiments. Scale bar= 5µm.

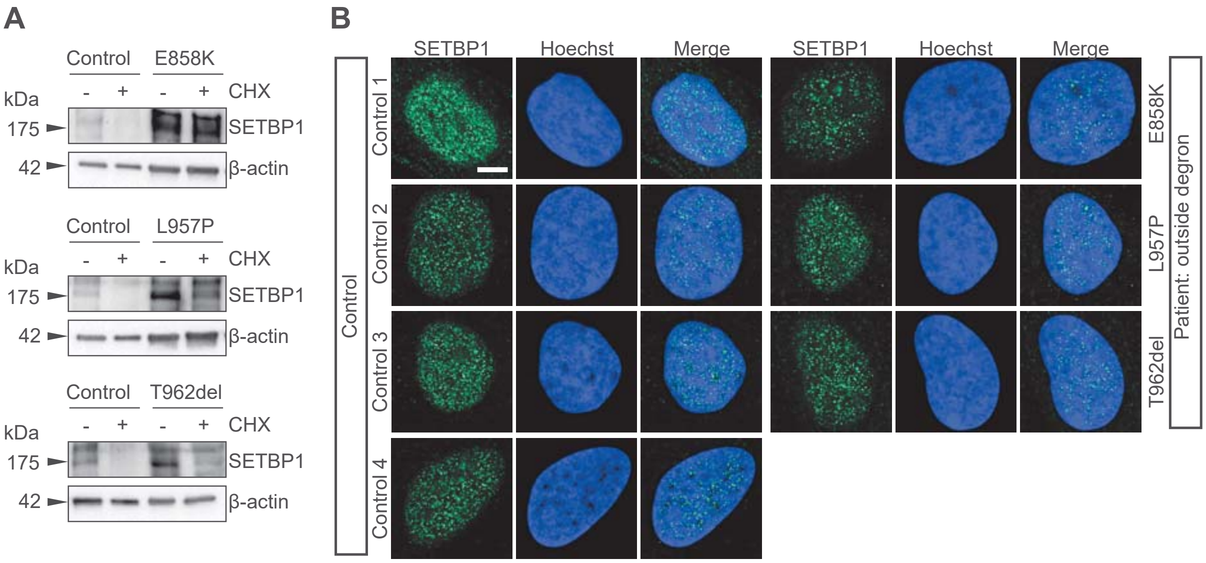

#### Figure S6: Relative expression of SETBP1 variants as YFP-fusion protein in HEK293T/17 cells treated with 50µg/mL cycloheximide (CHX) or DMSO vehicle control.

Normalized fluorescence intensity of YFP-SETBP1 in living HEK293T cells treated with 50µg/mL cycloheximide (CHX) or equal volume of DMSO as vehicle control. Fluorescence intensity was measured for 24 hours with three-hour intervals and normalised to the transfection control mCherry. Values are expressed relative to t= 0 hour and represent the mean ± SEM of three independent experiments, each performed in triplicate (*p<0.05, **p<0.01, ***p<0.001, ****p<0.0001 CHX versus DMSO; repeated measure two-way ANOVA and a post-hoc Sidak's test).

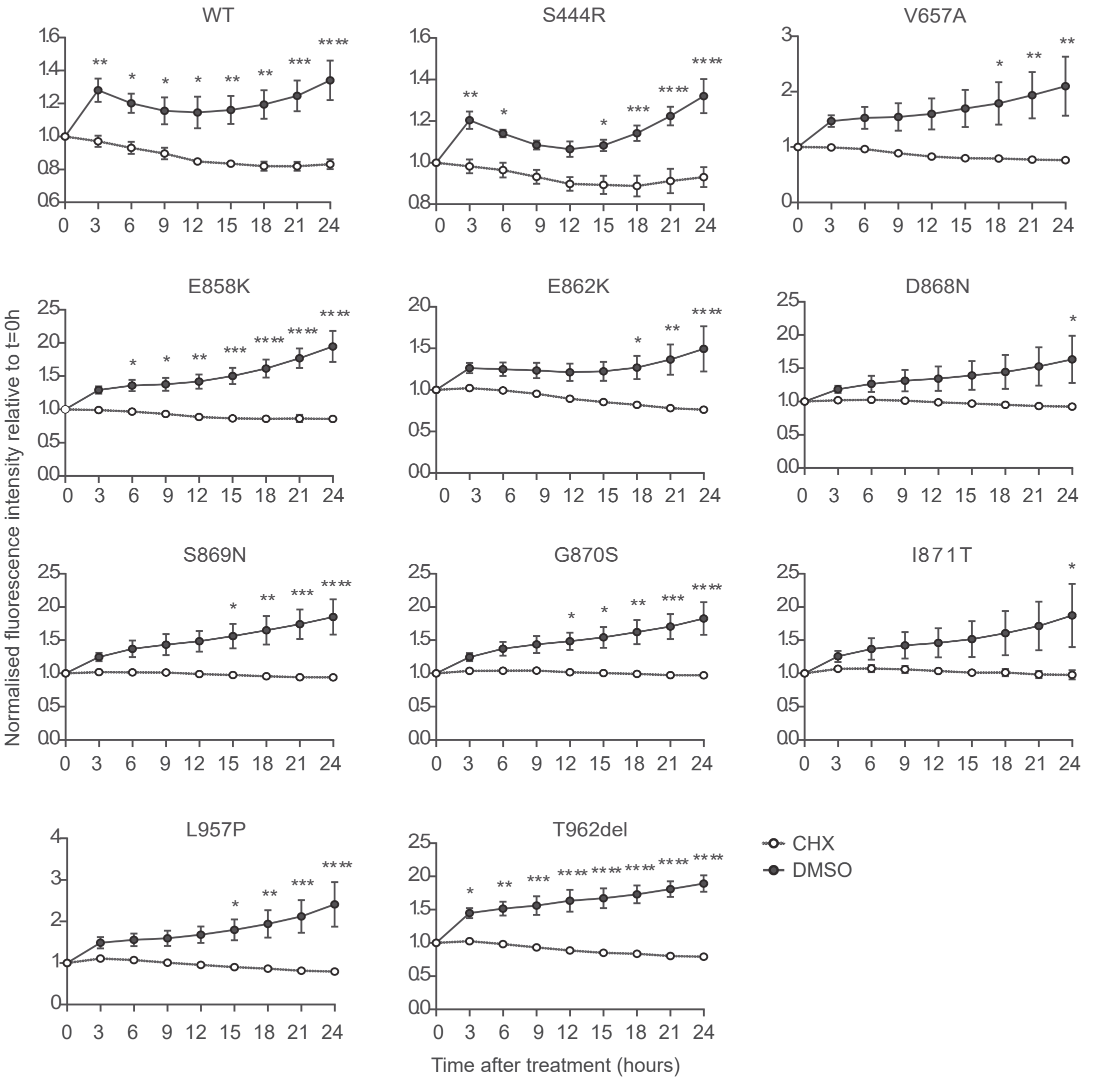

#### Figure S7: Relative expression of SETBP1 variants as YFP-fusion protein in HEK293T/17 cells treated with 5µg/mL MG132 or DMSO vehicle control.

Normalized fluorescence intensity of YFP-SETBP1 in living HEK293T cells treated with 5µg/mL MG132 or equal volume of DMSO as vehicle control. Fluorescence intensity was measured for 24 hours with three-hour intervals and normalised to transfection control mCherry. Values are expressed relative to t= 0 hour and represent the mean ± SEM of three independent experiments, each performed in triplicate (*p<0.05, **p<0.01, ***p<0.001, ****p<0.0001 MG132 versus DMSO; repeated measure two-way ANOVA and a post-hoc Sidak's test).

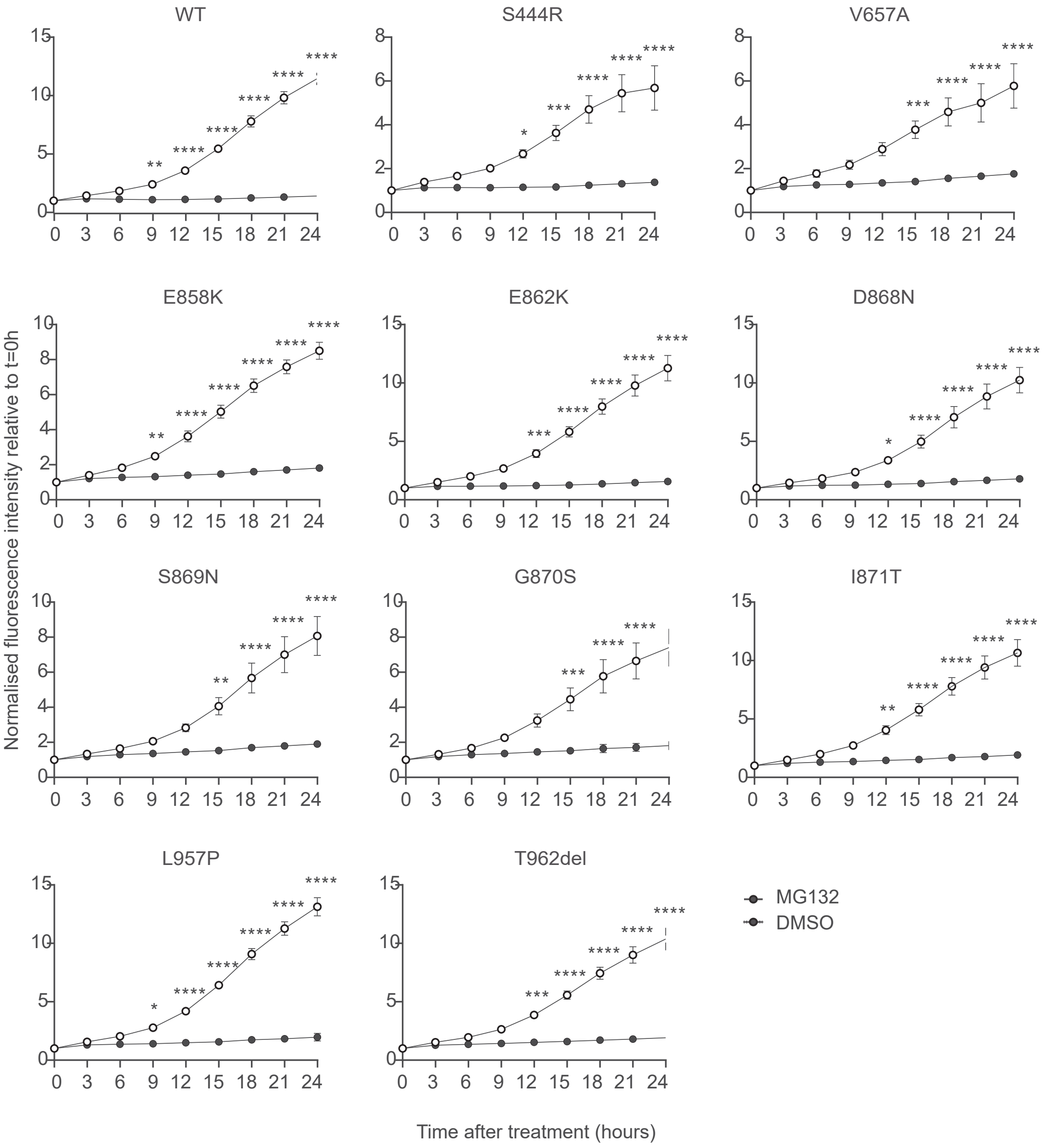

#### Figure S8: Relative expression of SETBP1 variants as YFP-fusion protein in HEK293T/17 cells treated with 100nM Bafilomycin (BafA1) or DMSO vehicle control.

Normalized fluorescence intensity of YFP-SETBP1 in living HEK293T cells treated with 100nM BafA1 or equal volume of DMSO as vehicle control. Fluorescence intensity was measured for 24 hours with three-hour intervals and normalised to transfection control mCherry. Values are expressed relative to t= 0 hour and represent the mean ± SEM of three independent experiments, each performed in triplicate (*p<0.05, **p<0.01, ***p<0.001, ****p<0.0001 BafA1 versus DMSO; repeated measure two-way ANOVA and a post-hoc Sidak's test).

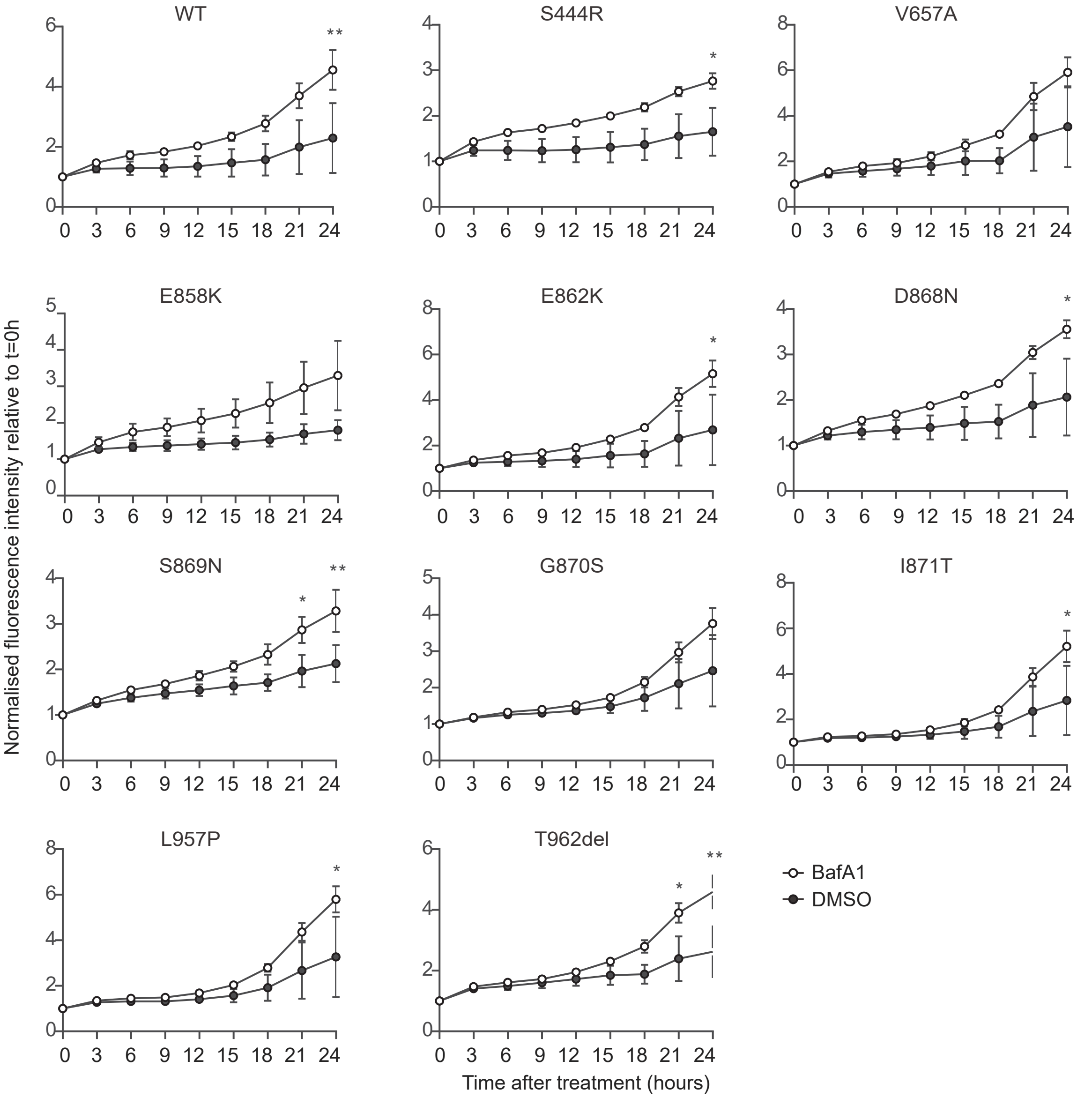

#### Figure S9: SETBP1 affinity to two AT-rich DNA consensus sequences.

(A) Expression constructs used in the luciferase reporter assays: a YFP-fused SETBP1 construct under a CMV promoter; a control construct with Renilla luciferase under control of a TK promoter for normalisation; and a promoter-less Firefly luciferase reporter construct carrying six repeats of 5'-AAAATAA-3' or 5'-AAAATAT-3' consensus SETBP1 binding sequence previously reported in Piazza *et al.*[1] (top panel). In the mammalian-one-hybrid (M1H) assay, a GAL4-fused SETBP1 construct with a CMV enhancer and promoter; a control construct with Renilla luciferase under control of a TK promoter; a Firefly luciferase reporter construct with or without a GAL4-binding site and an adenovirus major late promoter (AMLP) (middle panel). Expression constructs used in the luciferase reporter assays: a YFP-fused SETBP1 construct under a CMV promoter; a control construct with Renilla luciferase under control of a TK promoter for normalisation; and a promoter-less Firefly luciferase reporter construct carrying FOXP2 promoters (TSS1 or TSS2) previously reported in Becker *et al.*[2] (bottom panel). (B) Results of luciferase assays with pYFP constructs containing WT or SETBP1 variants, and the Firefly luciferase reporter constructs carrying six repeats of 5'-AAAATAA-3' or 5'-AAAATAT-3' consensus SETBP1 binding sequence previously reported in Piazza *et al.*[1]. Values are expressed relative to the control condition that used a pCMV-YFP construct without SETBP1 and represent the mean ± SEM of three independent experiments, each performed in triplicate (*p<0.05, student's t-test).

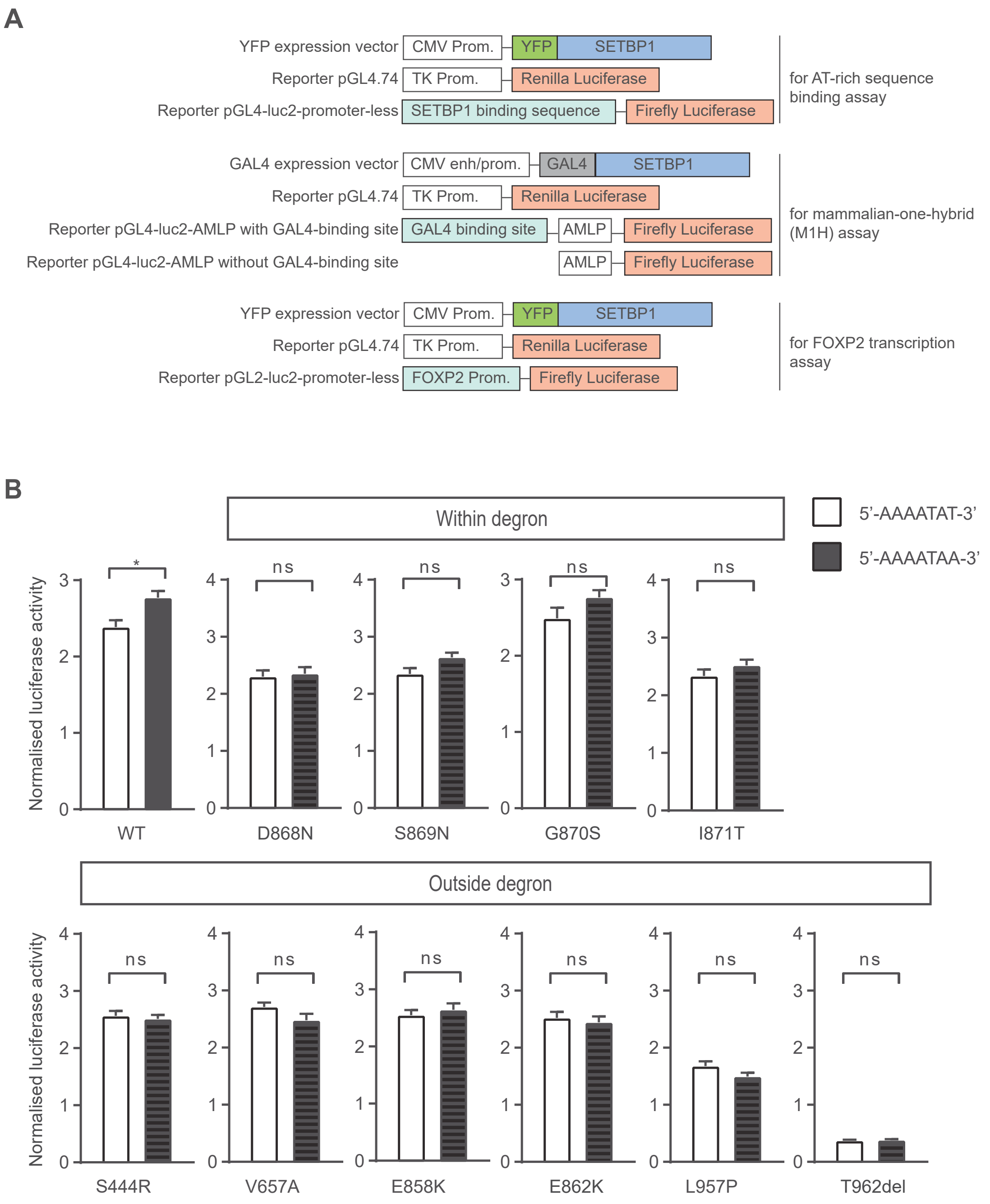

#### Figure S10: Proliferation of patient fibroblasts carrying SETBP1 variants outside the degron and SETBP1 interaction with SET.

(A) Confocal microscopy images of immunostained SETBP1 (green) and proliferation marker Ki67 (red) in control and patient fibroblasts. Nuclei were stained with Hoechst 33342 (blue). Merged images are shown. Ki67 shows a characteristic blob-like pattern in the nucleus in S phrase in all variants. Results are representative of three independent experiments. Scale bar= 5μ m. (B) Co-IP of GFP-SET in whole cell lysates co-expressing FLAG-SETBP1 and GFP-SET using GFP-trap or control beads and blotted with an anti-SET or anti-SETBP1 antibody. Results are representative of three independent experiments. (C) Normalised SET expression in control and patient fibroblasts. Bars represent the mean ±SEM of three independent experiments (not statistically significant vs controls; one-way ANOVA and a post-hoc Dunnett's test).

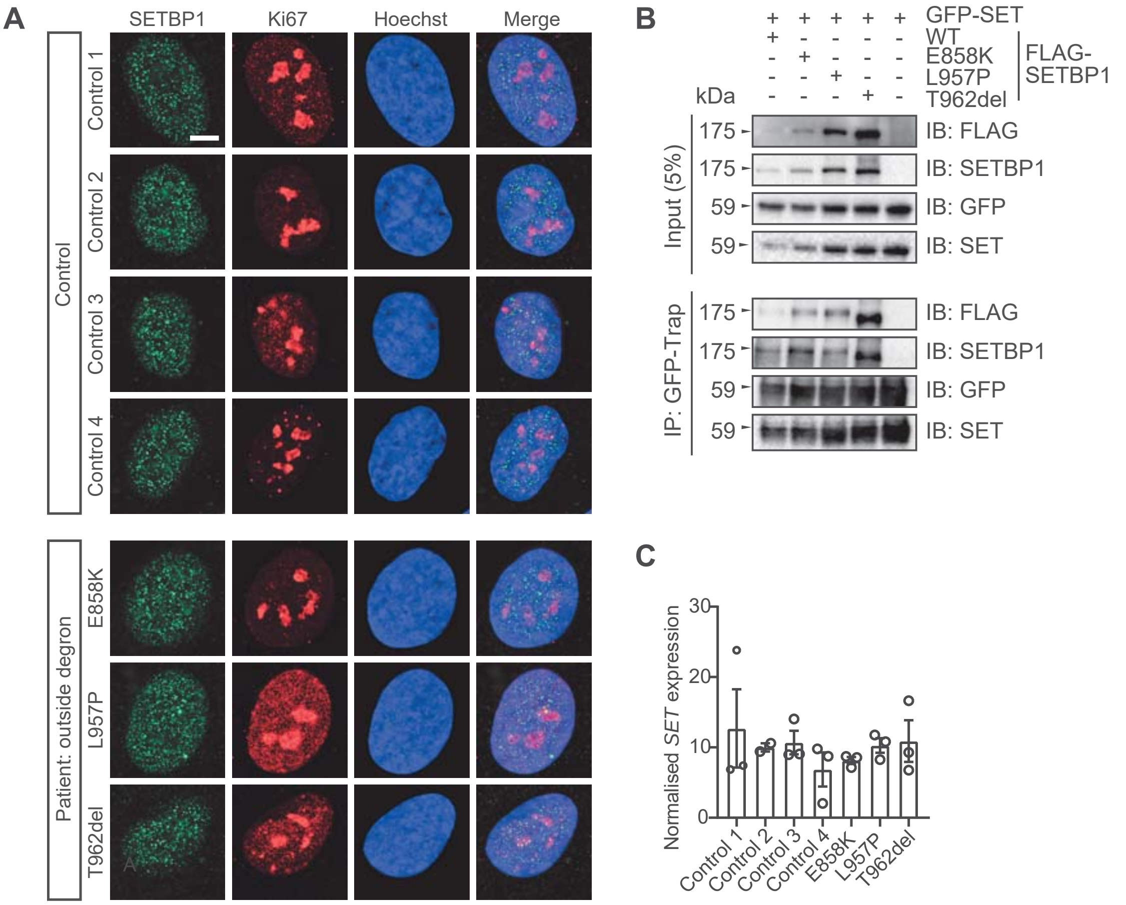

#### Figure S11: Venn diagrams and gene ontology analysis of differentially expressed genes in fibroblasts.

(A) Venn diagram of differentially expressed genes [DEGs (DESeq2 and intensity differences hits)] in patient human dermal fibroblasts (HDF) with variants outside the degron and genes associated with autism or intellectual disability (ID) in PanelApp. (B) Venn diagram of overlap between 32 DEGs in patient HDF carrying missense variants outside the degron in this study and previous reported DEGs in HEK293 cells overexpressing a SETBP1 SGS variant G870S versus empty vector[1]. Nine DEGs were also dysregulated in both data sets, suggesting partially overlapping disease pathways. (C) Venn diagram of 32 DEGs in patient HDF harbouring variants outside the degron and previously reported putative SETBP1 direct targets identified by ChIPseq[1]. RUNX3 was dysregulated in HDF harbouring SETBP1 variants and a putative direct target of SETBP1. (D) Venn diagram of previously reported putative SETBP1 direct targets identified by ChIPseq[1] and genes associated with autism or ID in PanelApp. Among those, 21 putative SETBP1 direct targets were associated with autism or ID. (E) Dysregulated GO biological process and cellular components revealed by over-representation analysis of the 32 DEGs in patient fibroblasts using gene set enrichment analysis (GSEA, $p<0.05$, multiple testing correction with FDR).

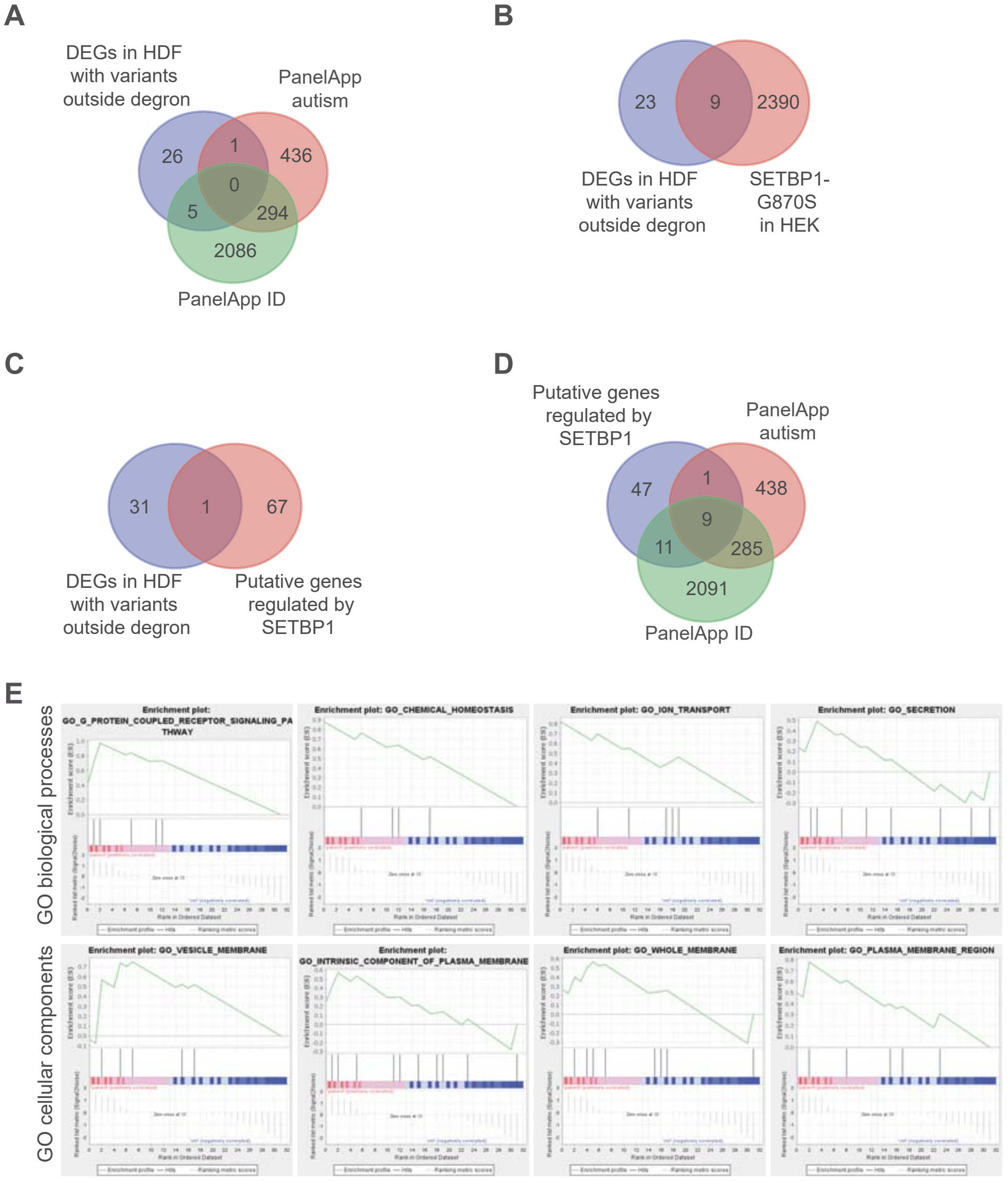
